# Microstructural Alterations in White Matter Hyperintensities and Perilesional Normal-Appearing White Matter Assessed by Quantitative Multiparametric Mapping - A BeLOVE Study

**DOI:** 10.64898/2026.04.10.26350576

**Authors:** Huma Fatima Ali, Markus G. Klammer, Tobias Leutritz, Ralf Mekle, Andrea Dell’Orco, Stefan Hetzer, Joachim E. Weber, Michael Ahmadi, Sophie K. Piper, Simrit Rattan, Katharina Schönrath, Ira Rohrpasser-Napierkowski, Nikolaus Weiskopf, Jeanette Schulz-Menger, Anja Hennemuth, Matthias Endres, Kersten Villringer, the BeLOVE-study group

**Author notes:** Corresponding author: Huma Fatima Ali, Address: Charité – Universitätsmedizin Berlin; Center for Stroke Research Berlin. Charité campus Benjamin Franklin, Hindenburgdamm 30, 12203, Berlin, Germany.

## Abstract

**Background and Objectives:** Normal appearing white matter (NAWM) may already harbor subtle microstructural alterations not yet visible on conventional MRI. Quantitative Multi-Parametric Mapping (qMPM) such as Magnetization Transfer saturation (MTsat), longitudinal relaxation rate (R1), and Proton Density (PD) offer new possibilities for analyzing NAWM which are sensitive to demyelination, axonal loss, and edema. We aimed to characterize these alterations within white matter hyperintensities (WMH) and the perilesional NAWM (pNAWM), to gain insights into the underlying process of lesion progression. We also investigated their association with cerebrovascular risk factors (CVRF) and long-term cognitive performance.

**Methods:** This investigation included the cerebral MRI data of 245 participants from the prospective Berlin Longterm Observation of Vascular Events (BeLOVE) study. Furthermore, 121 participants’ cognitive performance was evaluated at baseline and longitudinally at 2 years follow-up using Montreal Cognitive Assessment (MoCA). Regions of interest (ROIs) of WMH, pNAWM at 1, 2, 3 mm were assessed in comparison to the mirrored contralesional white matter (cWM). Linear mixed effects models were employed to demonstrate the pairwise comparisons between each region using estimated marginal means and the association of MPM metrics with CVRFs. Linear regression was used to assess the association with cognitive performance.

**Results:** In 245 participants, (mean age 62 years, SD: 12 years; 29.8% females), MPM metrics demonstrated a clear spatial gradient of microstructural injury. MTsat and R1 values were lower in WMH compared to cWM (ß = -0.48 (-0.52 - -0.44) and ß = -0.07 (-0.08 - -0.06), p<0.001, respectively) and showed gradual recovery with increasing distance indicating a microstructural gradient in pNAWM. Conversely, PD values were higher in WMH and decreased peripherally (ß = 2.32 (2.05 – 2.61, p<0.001). No substantial associations were found between MPM parameters and CVRFs in our cohort. At baseline and 2-year follow-up, cognitive performance was associated with higher pNAWM R1 values, whereas MTsat were only moderately associated.

**Discussion:** Quantitative MPM reliably detects microstructural alterations not only within WMH, but also in pNAWM, confirming the high sensitivity of qMPM to subtle tissue pathology and support its utility as a promising biomarker for longitudinal studies and monitoring therapeutic effects.

## Introduction

Cerebral small vessel disease (CSVD) represents one of the most prevalent neuropathological processes known to be associated with aging and vascular risk factors and is considered a major contributor to stroke, gait dysfunction, and cognitive decline^1,2^. Its radiological hallmarks are white matter hyperintensities (WMH), lacunes, microbleeds, and enlarged perivascular spaces, reflecting a spectrum of microvascular injury^3,4^. Of these biomarkers, WMH in particular has been associated with impaired cognitive performance, especially in domains of executive function, processing speed, and complex attention^5–7^. Although WMH volume provides a useful proxy for disease burden, accumulation of WMH represents a late manifestation of tissue injury^8,9^. Growing evidence indicates that substantial microstructural damage evolves within normal-appearing white matter (NAWM) long before lesions become radiologically visible^10,11^.

A wide range of cerebrovascular risk factors (CVRF) contribute to the development and progression of CSVD^12,13^. Age remains the strongest determinant of WMH burden, but hypertension, diabetes mellitus (DM), and obesity have each been linked to disrupted white matter integrity and poorer cognitive outcomes^13–15^. In addition, greater arterial stiffness has been found to be independently linked to increased WMH burden^16^. These associations reflect the systemic impact of metabolic and hemodynamic stress on the cerebral microvasculature, resulting in chronic hypoperfusion, endothelial dysfunction, blood–brain barrier leakage, and ultimately myelin disruption^17–19^. Although prior studies have demonstrated relationships between CVRFs and WMH volume^12,20^, less is known about the extent to which vascular risk factors influence the microstructure of NAWM in regions adjacent to WMH, where tissue may appear radiologically normal yet is histologically compromised^9–11^.

Advances in quantitative MRI have significantly improved the ability to detect microstructural white matter injury. Multiparametric mapping (MPM) provides high-resolution, quantitative measures of tissue microstructure through Magnetization Transfer saturation (MTsat), longitudinal relaxation rate (R1), and Proton Density (PD) maps^21–24^. Each map captures distinct biophysical properties and shows differential sensitivity to the underlying microstructure: MTsat and R1 are sensitive to macromolecular content and myelin-associated components. PD reflects water content and tissue density^25,26^. Thus, MT and R1 are frequently anti-correlated to PD^27^. Previous work has demonstrated that these quantitative measures detect subtle abnormalities in NAWM in CSVD, multiple sclerosis, and neurodegenerative disorders^10,28–30^. However, their regional behavior across WMH and perilesional NAWM (pNAWM), particularly in vascular cohorts, remains insufficiently characterized. Moreover, their relevance for cognitive function and their susceptibility to CVRF have not been comprehensively addressed.

Emerging evidence suggests that WMH are not isolated lesions but part of a spatially distributed process that extends into the surrounding NAWM^9,11,31–33^. Diffusion MRI studies have consistently shown microstructural gradients radiating outward from WMH, with abnormalities attenuating as distance increases^9,34,35^. Whether similar gradients are detectable using MPM metrics and whether such gradients carry information regarding cognitive performance has not yet been fully established. Given that cognitive impairment in CSVD cannot be explained by WMH volume alone, identifying sensitive markers of NAWM integrity may yield insights into early microvascular injury that contributes to cognitive decline^15,36^.

Against this background, the present study sought to provide a detailed, quantitative characterization of NAWM microstructure using MPM-derived MTsat, R1 and PD maps, evaluating microstructural properties across WMH, pNAWM at increasing spatial distances, and contralesional white matter (cWM). Furthermore, we examined the relationship between these microstructural markers and (1) a panel of CVRFs, and (2) cognitive performance at baseline and after two years of follow-up.

## Methods

### Cohort

This is an analysis of an ongoing prospective longitudinal observational cohort study Berlin Longterm Observation of Vascular Events (BeLOVE) conducted by the Berlin Institute of Health (BIH) at Charité Universitätsmedizin Berlin, Germany (DRKS00016852). This study recruits participants ≥18 years of age, within 0-7 days after a stroke/transient ischemic attack, myocardial infarction (MI), acute kidney injury, or acute heart failure, or with chronic vascular high-risk conditions, and performs a comprehensive deep phenotyping at 3 months and following years after the acute event or study inclusion in the chronic risk sub-cohort. Detailed inclusion and exclusion criteria of the study can be found in the previously published study protocol^37^. A subset of data acquired (BeLOVE phase 1.0) between 07/2017 to 12/2021 has been selected for this study with analyses performed on data extracted from database on 23/05/2024. The ethics committee approved the study for all recruiting centers in Berlin in accordance with the Declaration of Helsinki (EA1/066/17).

For this study, the 3 months deep phenotyping visit was defined as baseline. Patients with Age-Related White Matter Changes (ARWMC) score^38^ of ≤8 were included in the analysis while patients with ARWMC score of >8, presence of large infarcts covering >50% of the corresponding supplying artery, multiple infarcts especially in the white matter, and Hyperintense Acute Reperfusion Marker (HARM) sign on fluid-attenuated inversion recovery (FLAIR) sequences were excluded from analysis. Therefore, out of 503 participants of BeLOVE phase 1.0 with a cerebral MRI after 3 months, 245 were included for final analysis. Furthermore, cognitive performance was analyzed using Montreal Cognitive Assessment (MoCA) at baseline (n=173) and at follow-up of two years in a sub-cohort of 121 participants to investigate long-term effect of MPM metrics on cognitive decline (**Figure 1**).

**Figure 1:**
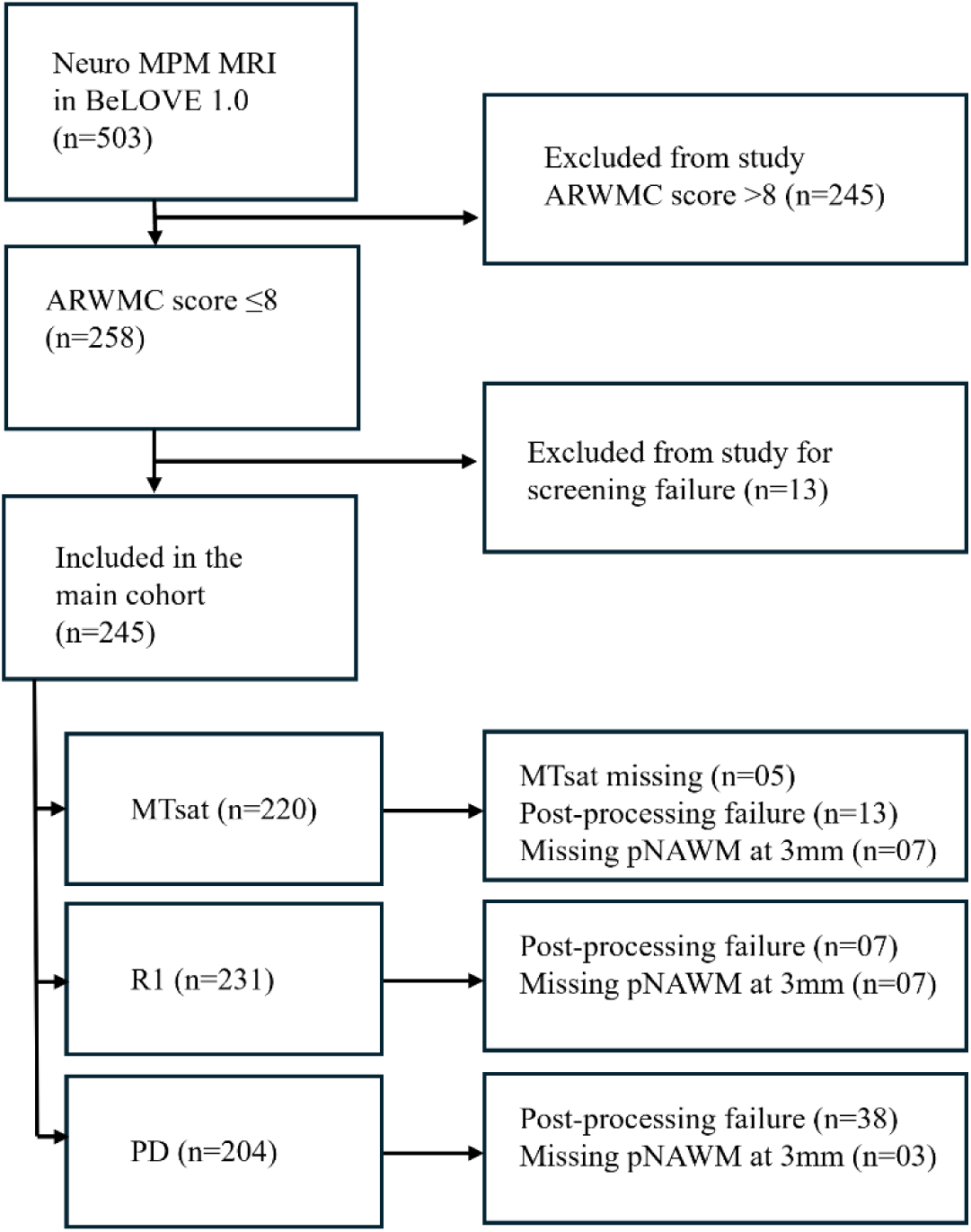
Patient selection flow chart. BeLOVE = Berlin Long term Observational of Vascular Events; MRI = Magnetic Resonance Imaging; ARWMC score = Age-Related White Matter Changes score; MTsat =Magnetization Transfer saturation map; R1 = Longitudinal relaxation rate map; PD = Proton Density map; MoCA = Montreal Cognitive Assessment.

### MRI assessment

MPM sequences were acquired using a Siemens 3T Tim Trio (upgraded to Prisma Fit 1-year after the study onset; Siemens Healthineers, Erlangen, Germany) at Charite Universitätsmedizin Berlin (campus Mitte and campus Benjamin Franklin), with longitudinal relaxation time (T1), proton density (PD), and magnetization transfer (MT) weighting using the following acquisition parameters: TR=18ms, TE 6 echoes (2.46-14.76), flip angle 25°, matrix 256x256, 1mm slice thickness, and voxel size of 1x1x1mm. For the MT-contrast, an off-resonant MT saturation preparation pulse (duration = 8ms, flip angle = 220°, frequency offset = 1000 Hz) was employed. Furthermore, the creation of MPM maps i.e., MTsat, R1, and PD was achieved using an automated pipeline developed by S.H. based on hMRI toolbox^23^ (https://hMRI.info) and pre-processed using SPM12 ((https://www.fil.ion.ucl.ac.uk/spm/software/spm12/)) which included Gibb’s ringing correction, parameter map quantification using ESTATICS model with B1+ bias field correction and normalization to ICBM template space of European brains.

WMHs were visually rated (HFA and KV) using the ARWMC scale on FLAIR sequences which ranges from a score of 0 to 30^38^. Only participants with ARWMC Score of ≤ 08^39^ were included in the analyses representing mild to moderate WMH lesion load to avoid confluent gliotic lesions.

Each included participant’s FLAIR and all three MPM maps were first co-registered to their structural T1weighted MPRAGE images individually to account for contrast differences, followed by their reorientation to the Montreal Neurological Institute (MNI) space. Brain extraction was performed on co-registered and reoriented FLAIR and a binary brain-extracted FLAIR mask was created. Using the binary FLAIR mask, brain extraction was performed on the MPM maps using FSL subtraction (https://fsl.fmrib.ox.ac.uk/fsl/docs/utilities/fslutils.html). A region of interest (ROI) was manually segmented on the post-processed FLAIRs by selecting a single punctate gliosis for each case having sufficient pNAWM and cWM. ROIs were segmented using MRIcron Software from the Center for Advanced Brain Imaging (University of South Carolina, Chris Rordan, USA; NITRC: MRIcron: Tool/Resource Info). Each WMH ROI was dilated by 1mm to cope with potential subtle coregistration mismatches between FLAIRs and MPM maps. These ROIs were mirrored onto the opposite hemisphere to create cWM ROIs. This was followed by the creation of pNAWM masks by dilating ROIs in increasing steps of 1 mm up to 3 mm for each participant, respectively, then subtracting each dilated ROI from the previous one using fslmaths. Special care was taken not to overlap with any other type of tissue, and in cases where 3 mm NAWM ROI exceeded NAWM and overlapping grey matter or ventricles, were corrected only to include NAWM. Cases with insufficient NAWM were excluded from analysis. (**Figure 2**).

**Figure 2:**
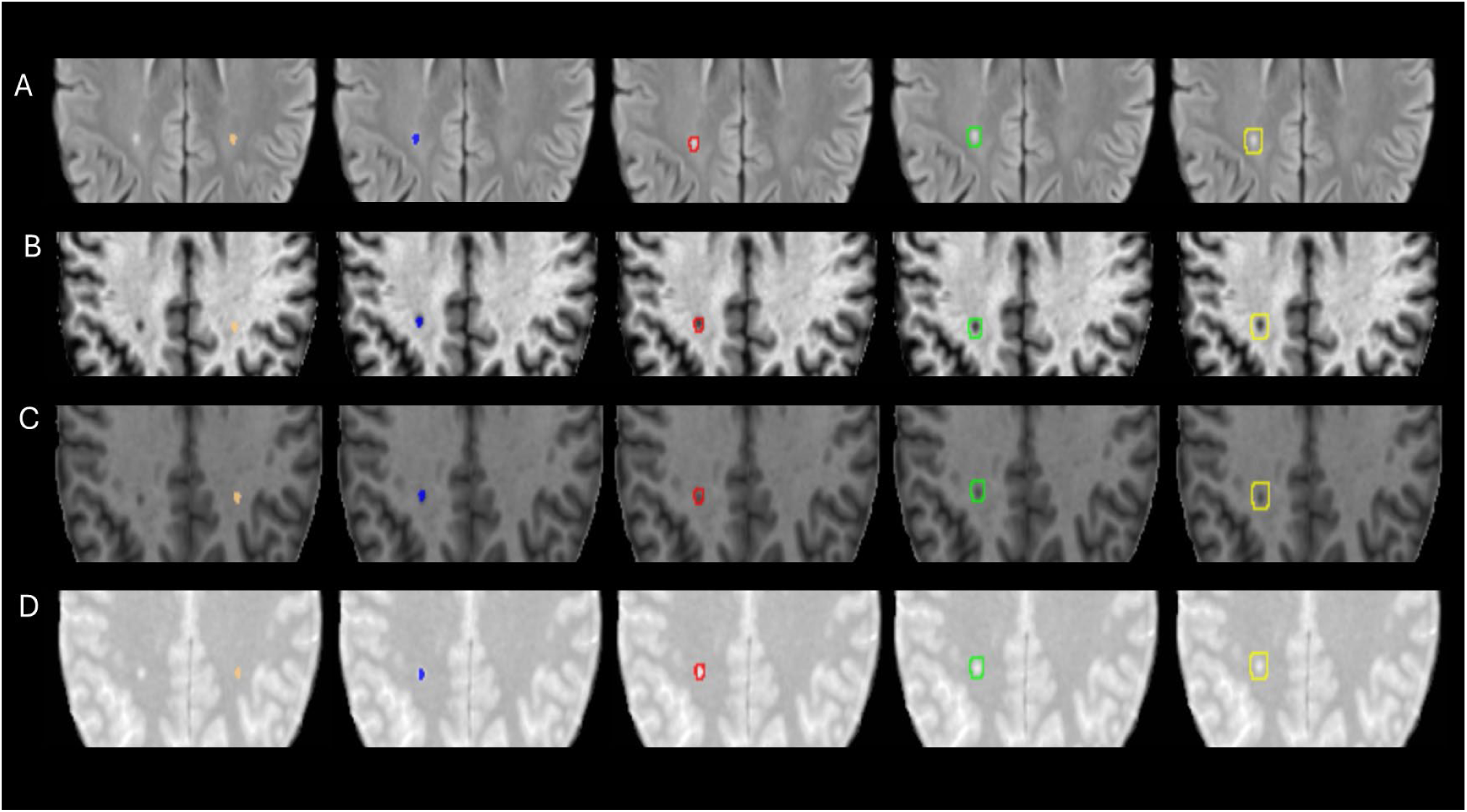
Multi-parametric maps (MPM) delineation framework. FLAIR (A) depicts segmented WMH (blue), cWM (beige), pNAWM at 1mm (red), 2mm (green) and 3mm (gold). Corresponding MTsat map (B), R1 map (C), and PD map (D) illustrate concentric ROI masks enabling voxel-wise quantification of microstructural gradients. ROI=regions of interest; FLAIR=Fluid attenuated inversion recovery; WMH=white matter hyperintensities; cWM=contralesional white matter; pNAWM=perilesional normal appearing white matter; MTsat= Magnetization transfer saturation; R1=longitudinal relaxation time; PD=proton density.

### Clinical and cerebrovascular risk assessment

Clinical parameters included basic patient demographics, education status, and CVRF profile and medication status. Pre-selected CVRF parameters were age, sex, DM, arterial hypertension, hyperlipoproteinemia, coronary artery disease, atrial fibrillation, smoking, waist-hip-ratio, Body Mass Index (BMI), pulse wave velocity (PWV), carotid intima-media thickness as well as serological markers (eGFR, serum creatinine, Interleukin-6, C-reactive protein, lipoprotein A, total cholesterol and Revised-Homeostatic Model Assessment (R-HOMA). These were documented and included into the analysis to study their association with microstructural alterations represented as MPM maps values.

### Cognitive assessment

Cognitive performance was assessed using the MoCA scale, a widely used screening tool for detecting cognitive impairment. The test yields a total score ranging from 0 to 30, with higher scores indicating better cognitive performance. A score below 26 is commonly used to indicate cognitive impairment in clinical and research settings^40^. Baseline scores assessed at 3-months visit and follow-up for two years were analyzed independently to investigate association between pNAWM microstructure and cognitive function.

### Statistical Analysis

Baseline characteristics of categorical data are represented in absolute and relative frequencies, the distribution of continuous data as mean ± standard deviation (SD) while scores and scales are represented as median and limits of the interquartile range (IQR).

Differences across white matter regions (cWM, WMH ROI, pNAWM at 1, 2, 3 mm) were evaluated using linear mixed-effects models with participant ID as a random intercept. This was done for MTsat, R1, and PD separately. Estimated marginal means were computed for each region. Pairwise contrasts were evaluated using Tukey-adjusted *p*-values to examine (1) deviation of each WMH and pNAWM region from cWM, and (2) stepwise changes in MPM metrics across increasing pNAWM distances of 1, 2, and 3mm.

To evaluate association between CVRFs and quantitative MPM metrics, linear mixed models with random intercept per patient were employed where separate models were fitted for each MPM map and each CVRF. The MPM values were the dependent variable and prespecified CVRF as independent variables. To assess whether associations between individual CVRF and MPM metrics varied across pNAWM distances from lesion borders, an interaction term (coded as MPM ∼ CVRF + region+CVRF*region +(1|ID)) was incorporated into the models. Potential pharmacological confounding was addressed through sensitivity analyses excluding participants receiving antihypertensive, antidiabetic, and/or lipid-lowering medications.

Associations between quantitative MPM metrics and cognitive performance were examined using linear regression and mixed-effects modeling approaches. Separate linear regression analyses were performed for MoCA scores at baseline and follow-up as dependent variables, respectively, and mean MPM metrics across pNAWM regions as independent variables. All models were adjusted for age, sex, and education status, while the follow-up models were additionally adjusted for baseline MoCA scores. Additionally, Spearman’s rank correlation coefficients were calculated to assess associations between mean R1 values in pNAWM at baseline and follow-up.

For all models, interpretation focused primarily on effect estimates and their 95% confidence intervals. A two-sided significance level of α = 0.050 was applied. However, since no adjustment for multiple testing was applied, p-values have to be interpreted cautiously. All analyses constitute exploratory data analyses and were performed in R (version 4.5.1 (2025.09.1+401); R Foundation for Statistical Computing) within the RStudio environment, using the lme4, emmeans, and stats packages. Graphical output and predicted probability curves were produced using ggplot2.

## Results

### Cohort characteristics

Baseline characteristics of the study population are summarized in **Table 1**. The mean (±SD) age was 62.2 ± 11.7 years, 73 (29.8 %) participants were females. The cohort exhibited a vascular risk profile with a prevalence of current smoking of 21.2 %, DM 40.0 %, arterial hypertension 80.8 %, history of heart failure 4.1 %, history of MI 6.1 %, and an incident stroke of 55.9 %. The WMH lesion load was moderate according to the inclusion criteria with ARWMC score median (IQR) of 4 (2-6).

**Table 1:**
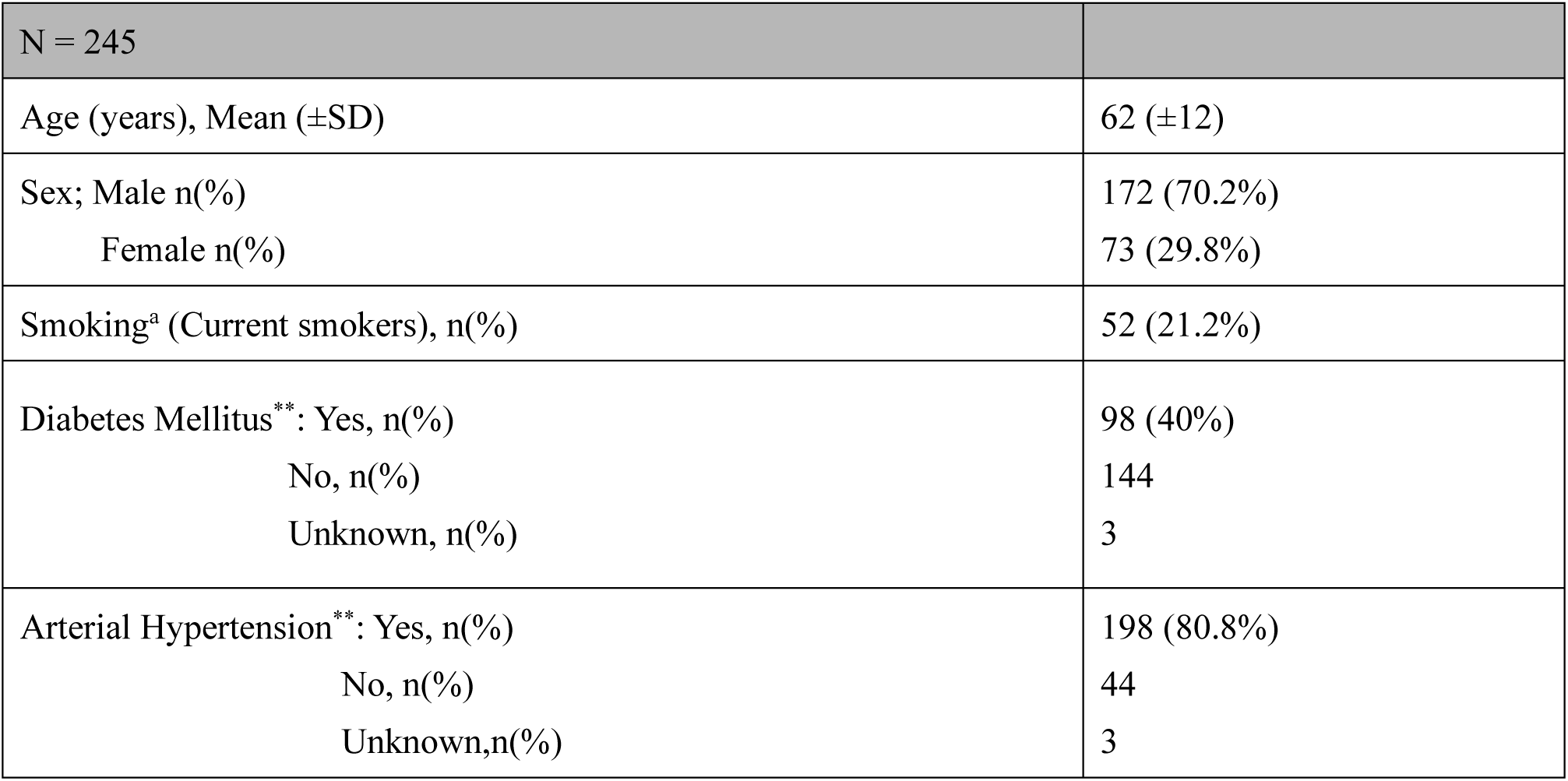
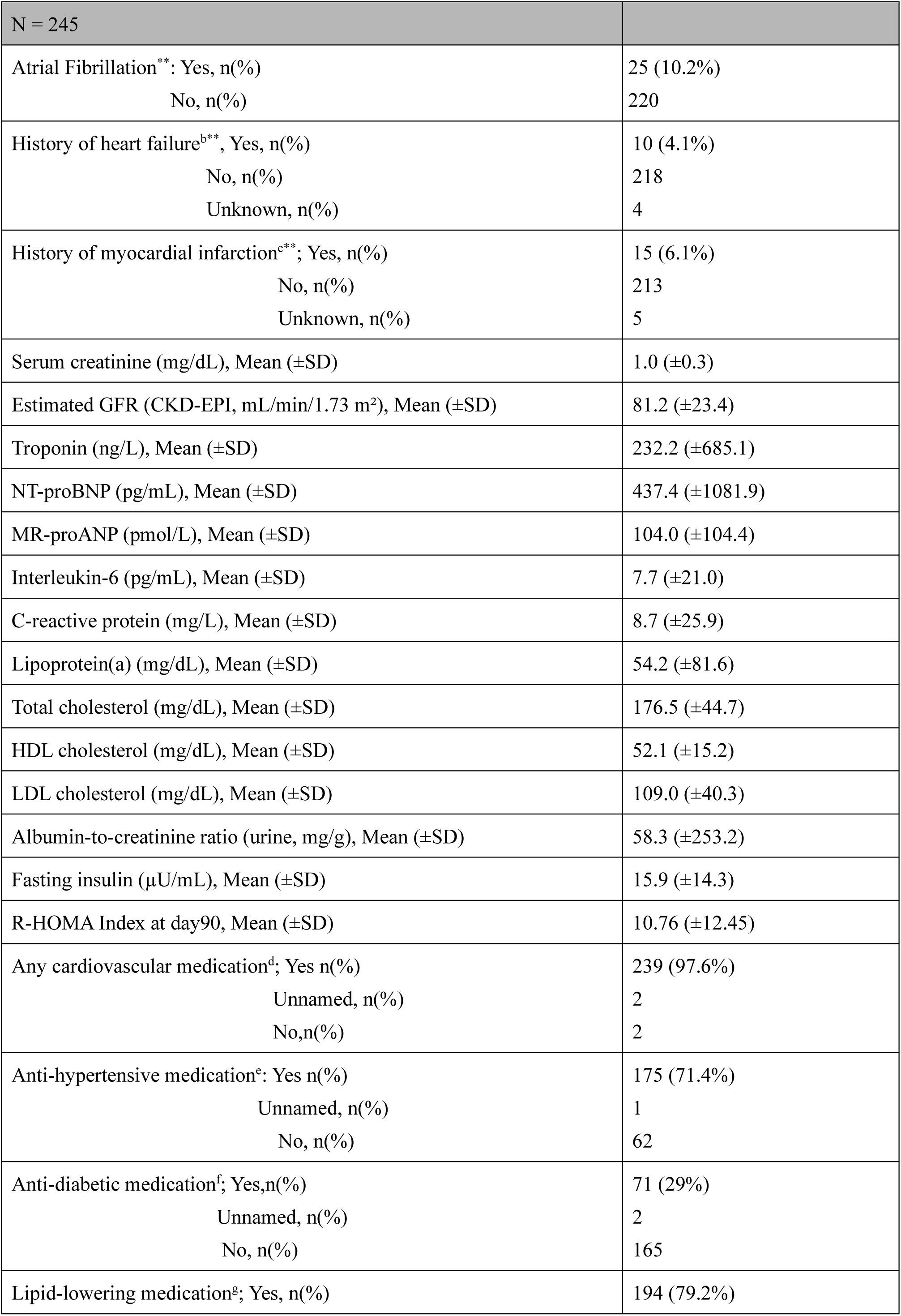
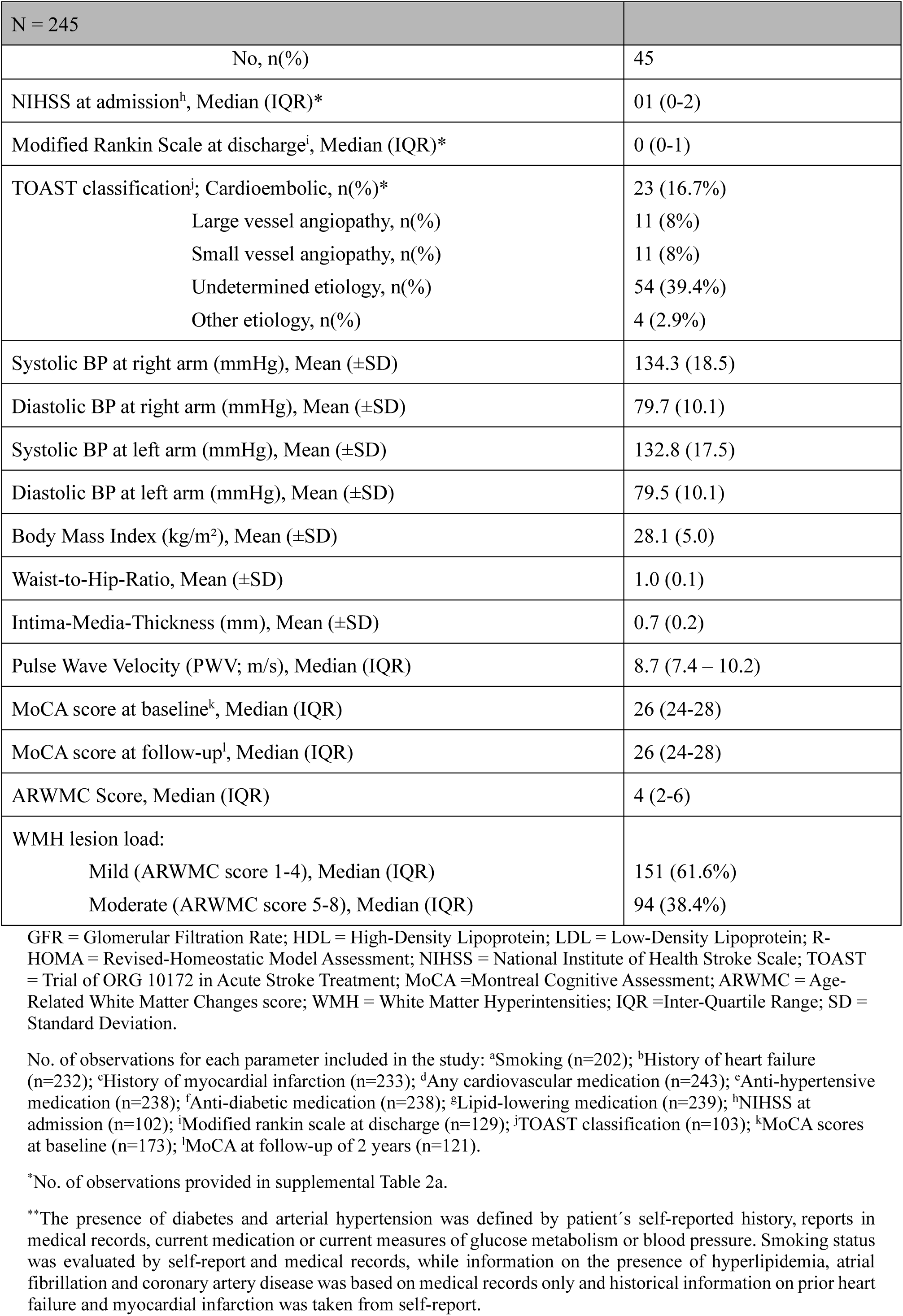
Baseline characteristics and clinical features of study cohort.

Global cognitive performance, as indexed by the MoCA, was relatively preserved at both time points, with a median (IQR) score of 26 (24–28) at baseline and an identical distribution at two-year follow-up. Additional stratification of demographic, vascular, and biochemical variables across BeLOVE subcohorts has been provided in **Supplementary Table 1**.

### Spatial pattern of NAWM microstructure across MPM maps

A quantitative assessment of white matter microstructure across the predefined ROIs and mirrored cWM revealed a coherent spatial gradient in all three MPM metrics. WMH ROI showed the most pronounced microstructural derangement, characterized by markedly reduced MTsat, R1 and increased PD relative to cWM, consistent with myelin loss, axonal disintegration, and elevated free water content (**Figure 3**, **Table 2**).

**Figure 3:**
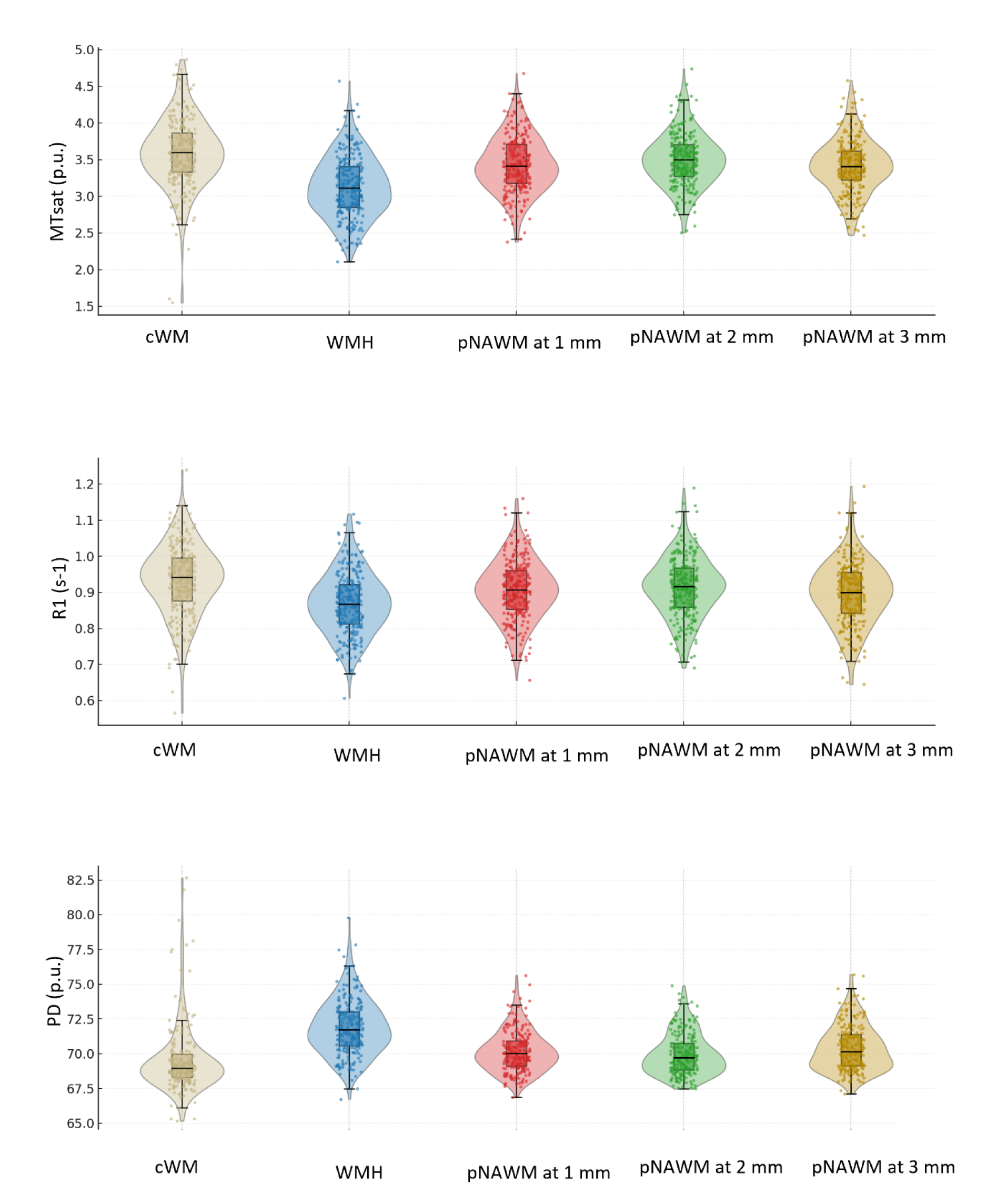
Quantitative MPM metrics across cWM (beige), WMH (blue), pNAWM at 1mm (red), pNAWM at 2mm (green), and pNAWM at 3mm (gold). MPM=Multi-parametric maps; WMH=White matter hyperintensities; cWM=contralesional white matter; pNAWM=perilesional normal appearing white matter; MTsat=Magnetization transfer saturation; R1=Longitudinal relaxation time; PD=Proton density; NAWM=normal appearing white matter.

**Table 2:** Linear mixed effects models for Spatial quantitative MPM microstructural metrics across white matter regions.

|  | MTsat (p.u.) |  | R1 (s<sup>-1</sup>) |  | PD (p.u.) |  |
| --- | --- | --- | --- | --- | --- | --- |
| Regions of interest | EMM (95% CI) |  | EMM (95% CI) |  | EMM (95% CI) |  |
| cWM | 3.61 (3.55 – 3.66) |  | 0.96 (0.92 – 0.99) |  | 69.44 (69.12 – 69.76) |  |
| WMH | 3.12 (3.07 – 3.18) |  | 0.89 (0.86 – 0.92) |  | 71.77 (71.45 – 72.09) |  |
| pNAWM at 1mm | 3.44 (3.39 – 3.49) |  | 0.93 (0.8 – 0.96) |  | 70.16 (69.84 – 70.48) |  |
| pNAWM at 2mm | 3.51 (3.45 – 3.56) |  | 0.94 (0.90 – 0.97) |  | 69.86 (69.54 – 70.18) |  |
| pNAWM at 3mm* | 3.43 (3.37 – 3.48) |  | 0.92 (0.89 – 0.95) |  | 70.31 (69.99 – 70.76) |  |
| Pairwise comparison with increasing distance | β (95% CI) | p-value | β (95% CI) | p-value | β (95% CI) | p-value |
| WMH and pNAWM at 1mm | -0.31 (-0.37 – -0.26) | <0.001 | -0.04 (-0.05 – -0.03) | <0.001 | 1.61 (1.22 – 2.0) | <0.001 |
| pNAWM at 1mm and pNAWM at 2mm | -0.07 (-0.12 – -0.01) | 0.007 | -0.01 (-0.02 – 0.00) | 0.373 | 0.30 (-0.09 – 0.69) | 0.226 |
| pNAWM at 2mm and pNAWM at 3mm | 0.08 (0.02 – 0.13) | 0.001 | 0.02 (0.01 – 0.03) | 0.001 | -0.45 (-0.84 – -0.05) | 0.017 |
| Pairwise comparison with cWM as reference | β (95% CI) | p-value | β (95% CI) | p-value | β (95% CI) | p-value |
| WMH | -0.48 (-0.52 – -0.44) | <0.001 | -0.07 (-0.08 – -0.06) | <0.001 | 2.32 (2.05 – 2.61) | <0.001 |
| pNAWM at 1mm | -0.17 (-0.21 – -0.13) | <0.001 | -0.03 (-0.04 – -0.02) | <0.001 | 0.72 (0.44 – 0.99) | <0.001 |
| pNAWM at 2mm | -0.10 (-0.14 – -0.06) | <0.001 | -0.02 (-0.03 – -0.01) | <0.001 | 0.42 (0.14 – 0.70) | 0.028 |
| pNAWM at 3mm | -0.18 (-0.22 – -0.14) | <0.001 | -0.04 (-0.05 – -0.03) | <0.001 | 0.87 (0.58 – 1.15) | <0.001 |

MTsat = Magnetization Transfer saturation map; R1 = Longitudinal relaxation rate map; PD = Proton Density map; cWM = contralesional white matter; WMH = white matter hyperintensities; pNAWM = perilesional normal appearing white matter; p.u. percent units (MTsat and PD maps measuring units); s<sup>-1</sup> = measuring unit for R1 map.
\* pNAWM at 3mm missing observations: MTsat = 7; R1 = 7; PD = 3.

MTsat values were lowest within WMH ROI and increased spatially with distance into pNAWM. Compared to cWM, MTsat was lower in WMH (β = -0.48 (-0.52 - -0.44), p < 0.001), pNAWM at 1 mm (β = -0.17 (-0.21 – -0.13), p < 0.001), 2 mm (β = -0.10 (-0.1 4 – -0.06), p < 0.001), and 3 mm (β = -0.18 (-0.22 – -0.14), p < 0.001). Pairwise comparisons further demonstrated this spatial gradient with WMH ROI showing markedly lower MTsat than pNAWM at 1 mm (β = −0.31 (-0.37 - -0.26), p < 0.001), pNAWM at 1 mm lower as compared to 2 mm (β = −0.07 (-0.12 - -0.01), p = 0.007), and pNAWM at 2 mm was slightly higher than at 3 mm (β = 0.08 (0.02 – 0.13), p = 0.001).

R1 showed a similar but more subtle spatial pattern. WMH ROIs had lower R1 than cWM (β = -0.07 (-0.08 – -0.06), p < 0.001). The pNAWM at 1 mm (β = -0.03 (-0.04 – -0.02), p < 0.001), 2 mm ((β = -0.02 (-0.03 – -0.01), p <0.001) and 3 mm (β = -0.04 (-0.05 – -0.03), p < 0.001) all differed from cWM. Adjacent contrasts confirmed lower R1 in WMH ROI versus pNAWM at 1 mm (β = −0.04 (-0.05 - -0.03), p < 0.001), as well as between pNAWM at 2 mm and 3 mm (ß =0.02 (0.01 – 0.03), p = 0.001) .

PD demonstrated an inverted pattern relative to MTsat and R1, reflecting its sensitivity to tissue water content. PD was higher in WMH ROI compared with cWM (β = 2.33 (2.05 - 2.61), p < 0.001) and remained higher in pNAWM at 1 mm (β = 0.72 (0.44 - 0.99), p < 0.001), 2 mm (β = 0.42 (0.14 - 0.70), p = 0.028), and 3 mm (β = 0.87 (0.58 - 1.15), p < 0.001). Pairwise contrasts confirmed higher PD in WMH ROI than in pNAWM at 1 mm (ß = 1.61 (1.22 – 2.0), p < 0.001), and pNAWM at 3 mm showed higher PD relative to 2 mm (ß = -0.45 (-0.84 - -0.05), p < 0.017).

### Cerebrovascular risk factors and white matter microstructure

Across the three MPM maps, only a subset of CVRFs exhibited global associations with white matter microstructure, and only MTsat and R1 map showed associations within the pNAWM (**Supplementary Table. 2a**)

For MTsat, older age was associated with lower MTsat within WMH (β = −0.015 (-0.019 – -0.011), p < 0.001) reaching significane at pNAWM at 3mm (ß= 0.003 (0.000 – 0.006), p = 0.008), while waist-hip-ratio with pNAWM at 3mm (ß = 0.357 (-0.685 - -0.031), p = 0.032), BMI with pNAWM at 3mm (ß = -0.009 (-0.015 - -0.003), p = 0.002), and R-HOMA index with pNAWM at 3mm (β = -0.017 (-0.026 – -0.009), p = <0.001).

Lower R1 values were associated with higher waist-hip-ratio having main effect (ß = - 0.404 (-0.727 - -0.082), p =0.014) and at pNAWM at 3mm (ß = -0.118 (-0.187 - -0.049) p <0.001), while BMI showed regional association at pNAWM at 3mm (ß = -0.002 (-0.003 - - 0.000) p = 0.005), and R-HOMA-index as well at pNAWM at 3mm (ß =-0.003 (-0.004 - -0.000) p = 0.003).

Importantly, none of the examined CVRFs demonstrated substantial interactions with distance from the lesion and MPM metrics, indicating that their influence on white matter microstructure is predominantly diffused rather than focally accentuated in the immediate lesion border zone. Sensitivity analyses restricted to participants not receiving antihypertensive, antidiabetic or lipid-lowering medications yielded no substantial associations, suggesting that the observed relationships were not driven by pharmacological modulation (**Supplementary Table 2b**).

### NAWM microstructure and cognitive performance

At baseline, cognitive performance demonstrated selective associations with pNAWM microstructural integrity. Among all three MPM maps, R1 showed a positive association with MoCA scores (β = 1.457 (0.239 – 2.675), *p* = 0.019). MTsat and PD demonstrated no association with cognition (**Table 3**). While R1 related to baseline cognitive performance, it also showed significant association to cognitive performance with two-year follow-up (ß = 1.575 (0.037 – 3.113), p = 0.045) (**Table 3**). Moreover, while MTsat did not show any effect on cognitive performance at baseline but was found to be moderately associated at follow-up (β = 1.183 (-0.334 – 2.7), p =0.124).

**Table 3:**
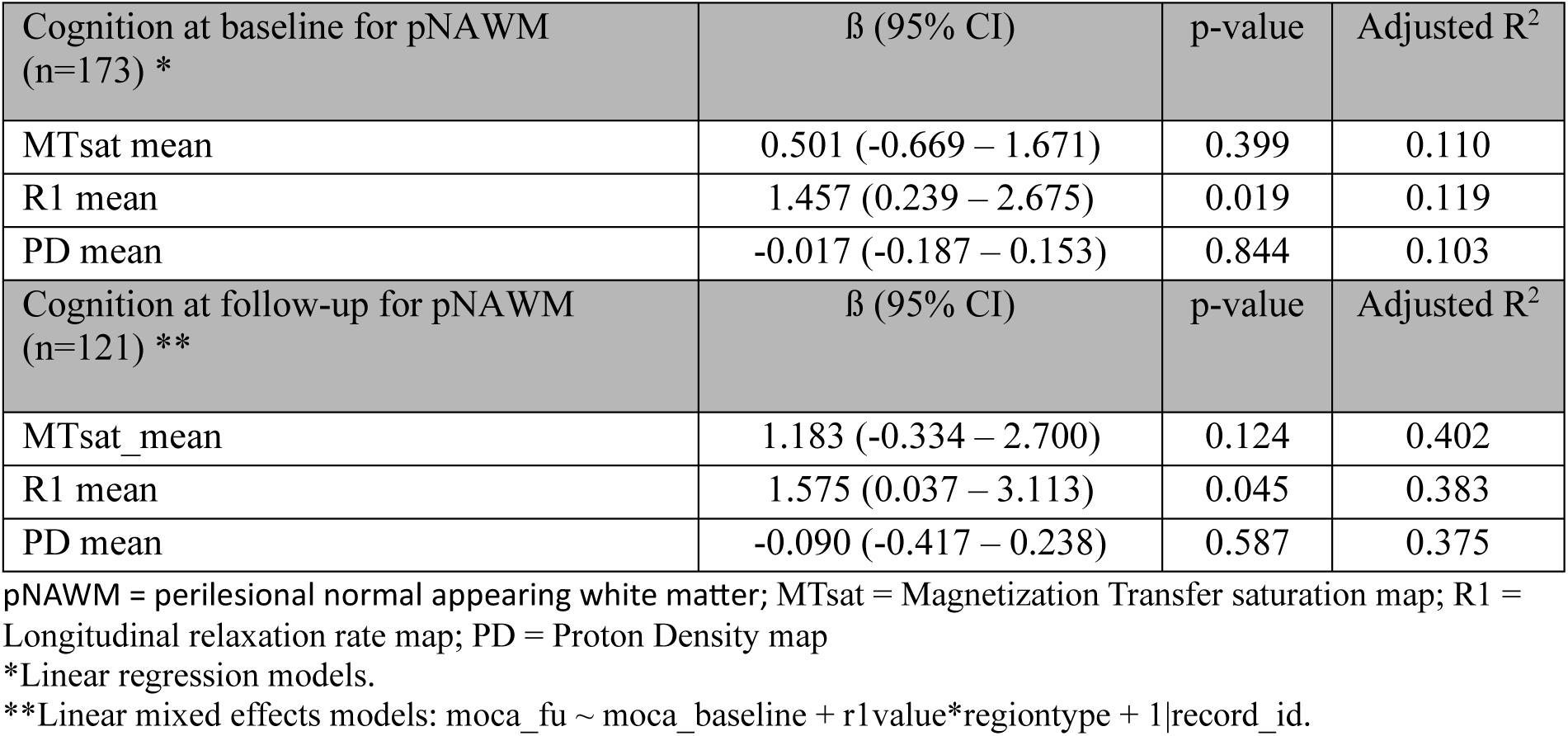
Association between NAWM MPM metrics and cognitive performance at baseline and follow-up.

| Cognition at baseline for pNAWM (n=173) * | $\beta$ (95% CI) | p-value | Adjusted R<sup>2</sup> |
| --- | --- | --- | --- |
| MTsat mean | 0.501 (-0.669 – 1.671) | 0.399 | 0.110 |
| R1 mean | 1.457 (0.239 – 2.675) | 0.019 | 0.119 |
| PD mean | -0.017 (-0.187 – 0.153) | 0.844 | 0.103 |
| Cognition at follow-up for pNAWM (n=121) ** | $\beta$ (95% CI) | p-value | Adjusted R<sup>2</sup> |
| MTsat_mean | 1.183 (-0.334 – 2.700) | 0.124 | 0.402 |
| R1 mean | 1.575 (0.037 – 3.113) | 0.045 | 0.383 |
| PD mean | -0.090 (-0.417 – 0.238) | 0.587 | 0.375 |

pNAWM = perilesional normal appearing white matter; MTsat = Magnetization Transfer saturation map; R1 = Longitudinal relaxation rate map; PD = Proton Density map
\*Linear regression models.
\*\*Linear mixed effects models: $moca\_fu \sim moca\_baseline + r1value*regiontype + 1|record\_id$ .

Spearman correlation analysis demonstrated a weak positive association between R1 values and baseline MoCA scores (ρ = 0.16, p = 0.040), and a stronger moderate association at follow-up (ρ = 0.34, p < 0.001) (**Supplementary Figure 1**).

## DISCUSSION

This study applied quantitative MPM to characterize the microstructural properties of white matter across radiologically identifiable WMH, adjacent pNAWM, and cWM in a mixed cohort of individuals with moderate to high cardio- and cerebrovascular risk. By integrating MPM metrics with cognitive performance and CVRF profiles, we identified microstructural signatures that extend well beyond visually identifiable WMH and demonstrated their relevance for short- and long-term cognitive performance.

Three principal observations emerged. First, there was a coherent spatial gradient of microstructural injury across all three MPM maps from gliosis into pNAWM. Second, pNAWM microstructural integrity, particularly as indexed by R1, showed meaningful associations with cognitive performance at baseline and over two-year follow-up. Third, while CVRFs exerted clear global effects on white matter microstructure, age, waist-hip-ratio, BMI and R-HOMA index demonstrated spatially region-specific effects with MTsat and some factors with R1 while PD demonstrated no region-specific effects within pNAWM. Collectively, these findings demonstrate the potential of MPM as a sensitive framework for detecting subvisual microstructural alterations in CSVD and for linking such changes to clinically relevant cognitive outcomes.

### Spatial gradients in microstructural integrity

The most salient finding was a spatially ordered microstructural gradient extending from WMH into the pNAWM assessed through MTsat, R1, and PD maps. This pattern of changes closely parallels histopathology of WMH, which emphasize demyelination, gliosis, and extracellular fluid accumulation^4,19^. Importantly, pNAWM MPM metrics indicated that microstructural degradation are not only confined to regions visible as hyperintense on FLAIR but radiates into tissue microstructure that appears normal on conventional imaging, creating a ‘penumbra’ of microstructural compromise in the pNAWM representing an intermediate state between the lesion core and healthy tissue^9,10,34^. This study establishes that these contrasts indicate a continuous perilesional gradient of partial microstructural recovery extending beyond the radiologically visible WMH boundary.

Similar spatial gradients in tissue integrity have been reported using diffusion MRI and myelin sensitive imaging^13,32^, consolidating current results by demonstrating a comparable pattern across three independent MPM metrics within a single quantitative framework. The behavior of the individual maps offers mechanistic insight. MTsat and R1, both sensitive to neuronal fiber integrity and myelin-related components, showed parallel gradients with marked reductions within WMH ROI and partial recovery with increasing distance from the lesion border. PD, which is inversely related to tissue density and sensitive to free water, exhibited an opposing gradient, with the highest values in WMH ROI and progressive normalization toward cWM values. These findings indicate persistently elevated free-water content and tissue degradation extending from WMH into the surrounding pNAWM, with a graded decline in PD as distance from the lesion increases. This pattern is in keeping with increased extracellular water content and reduced tissue fraction in pNAWM in only those patients exhibiting a CSVD score of >1 pointing to early pathogenesis of the disease^41^.

The central theme emerging from longitudinal studies is that WMH formation is not an acute event but rather the culmination of a continuous disease process characterized by the gradual loss of microstructural integrity in the pNAWM^31,42^. Furthermore, research utilizing Diffusion-Tensor Imaging (DTI) provides a foundational understanding of the pre-lesional stage establishing that pNAWM destined to convert into WMH already exhibits significantly altered microstructural properties years before becoming visible on conventional MR^8,19,43^. The robust differences between cWM and both WMH and pNAWM further suggest that even macroscopically normal white matter can harbor subtle microstructural abnormalities, in line with findings from MT and diffusion-imaging-based studies^10^. In this context, cWM is best considered a relatively “less affected” reference rather than truly healthy tissue, especially in patients with diffuse CSVD.

Taken together, MTsat, R1, and PD consistently demonstrated a continuum of white matter degradation extending beyond visibly WMH lesions (**Figure 3**). WMH occupied the most severely affected end of this spectrum, while pNAWM demonstrated graded microstructural alterations that partially, but not completely, normalized with increasing distance. The reproducible differences between cWM, WMH ROI and pNAWM suggest that even radiologically normal appearing tissue harbors a detectable microstructural signature of CSVD.

### NAWM microstructure and cognition

Among the MPM indices, R1 emerged as the most informative marker of tissue integrity in relation to cognition. Higher R1 values were associated with better MoCA scores at both baseline and follow-up, and this association was spatially consistent across pNAWM distances. These findings resonate with prior work showing that quantitative T1 and myelin-sensitive metrics capture subtle microstructural alterations that are cognitively relevant beyond the burden of overt WMH^44,45^. Lopes et al. demonstrated that WMH-derived tract-based disconnection factor showed strong association with cognitive impairment at 6- and 36-months post-stroke, independent of infarct-based features^46^. Although this study relied on lesion masks, current study demonstrates that quantitative MPM can refine these predictive models by providing a measure of tissue’s microstructural health potentially identifying pathological changes before functional outcome is affected.

MTsat showed weaker and less consistent associations with cognitive performance compared to R1, and PD exhibited minimal sensitivity to cognitive performance. This differential pattern reflects broader biophysical sensitivity of R1, which represents iron, myelin, and water effects^28^, whereas PD is predominantly driven by water content. Although MTsat specifically represents macromolecular content, its lower signal-to-noise ratio may have affected the association with cognitive scores in our cohort. Nonetheless, PD maps reflecting free water content appear to be less critical for global cognitive processing than the myelin-sensitive association assessed by R1 maps.

Earlier studies have established that higher WMH lesion load (ARWMC ≥ 10) increased the odds of cognitive decline at three years post-stroke^39^. Although WMH was quantified as a visual score, the study highlighted that macrostructural lesion burden remains a robust predictor of long-term cognitive performance, thereby motivating the search for more sensitive microstructural markers. With the current study, although R1 was associated with cognitive performance, other maps did not robustly predict the probability of cognitive decline over the short two-year interval. Nonetheless, the R1 analyses suggested a microstructural spatial gradient around MoCA score, wherein higher R1 in pNAWM was associated with a higher likelihood of cognitive performance (**Table 3**). This pattern echoes diffusion MRI studies that have linked white matter integrity to both global cognition and domain specific functions^47^ but adds a novel quantitative dimension by identifying R1 as a positive predictor of cognitive performance rather than merely a marker of damage.

### Cerebrovascular risk factors and microstructural integrity

Several CVRFs showed significant global associations with MPM metrics, most prominently age, waist-hip-ratio, BMI, and RHOMA-index. Age-related declines in MTsat are well documented and represent the known histological changes at microstructural levels^48^. The associations of ageand BMI with are compatible with microvascular injury, blood–brain barrier dysfunction and altered water homeostasis in these conditions^49,50^. Strikingly, however, few of CVRFs exhibited region specific associations within pNAWM predominantly with MTsat and R1 metrics. While PD showed no significant association with pNAWM regions and any CVRF, it is hence implied that rather than preferentially targeting tissue immediately adjacent to WMH, systemic risk burdens appeared to exert diffuse effects across the white matter (**Supplementary Table 2a).** This finding diverges from prior work that has emphasized associations between CVRFs and WMH volume or spatial distribution^5,19^. Nonetheless, these findings extends the research suggesting that pNAWM alterations may index global microvascular health rather than purely local vulnerability^48^.

Sensitivity analyses excluding individuals on medications did not modify these findings, suggesting that pharmacological treatment did not simply obscure underlying risk–microstructure relationships. Collectively, these results imply that pNAWM microstructural integrity, as assessed by quantitative MPM, is shaped more by intrinsic tissue vulnerability and global disease mechanisms than by regionally specific effects of conventional CVRF or medication status.

### Limitations and future directions

Several limitations of this study warrant consideration. The cross-sectional nature of this analysis represents a primary limitation, preventing causal inference regarding the temporal evolution of pNAWM. Consequently, it remains uncertain whether the observed microstructural gradients reflect the progressive spread of pathology from WMH into pNAWM, or preceding focal lesion formation, or both. Additionally, relying on cWM as a reference may lead to underestimated subclinical microstructural abnormalities due to the diffuse nature of CSVD, resulting in conservative effect sizes for lesion-non-lesional contrasts. Furthermore, the limited two-year cognitive follow-up period may be insufficient to fully characterize long-term cognitive trajectories.

Despite these constraints, the study provides strengths, particularly the quantitative characterization of pNAWM across spatially defined perilesional zones, a depiction not established with standard imaging techniques. The integration of both baseline and longitudinal cognitive measures reinforces the clinical relevance of MPM-derived markers. Moreover, the inclusion of a mixed cardiovascular cohort with inclusion of 30% females only, encompassing patients with CSVD and ischemic stroke, improves the generalizability and validity of the findings in high-risk populations. This has been noted that there is no simple one-to-one relationship between MPM parameters and the underlying microstructure and its alterations, while MPM metrics demonstrating preferential sensitity^24^. However, the use of multiple contrasts offering different perspectives on the underlying microstructure helps us disentangle and identify effects more clearly.

Future work should prioritize longitudinal MPM designs to track microstructural changes over time, with particular interest in the transition from pNAWM to overt WMH and in the dynamics of neuronal fiber disintegrity, demyelination, and free water accumulation. Multimodal imaging protocols, i.e., coupling MPM with other advanced modalities, such as DTI, may further clarify the vascular and inflammatory mechanisms underpinning white matter damage creating a composite microstructural-connectivity index. Moreover, incorporating MPM into interventional studies will be crucial to evaluate whether these metrics can serve as sensitive biomarkers for monitoring therapeutic response and predicting long-term cognitive outcomes.

## Conclusions

This study provides compelling evidence that quantitative MPM captures spatial microstructural gradients extending from WMH into pNAWM and cWM, demonstrating that pNAWM is not a healthy by-stander but an active site of continuous pathological processes that extend in graded manner from the lesion boundary. Among the MPM metrics, R1 emerged as the most sensitive indicator of tissue integrity linked to cognitive performance. Although systemic CVRF influences global white matter microstructure, they did not selectively influence pNAWM. These findings support quantitative MRI as a promising tool in assessing tissue pathophysiology in CSVD.

## Data availability statement

The data supporting the findings of this study are available from the corresponding author upon reasonable request and according to sharing agreements with the Berlin Institute of Health and Center for Stroke Research Berlin. Due to the presence of potentially identifying or sensitive patient information, source imaging data cannot be made publicly available due to participant privacy restrictions under the European General Data Protection Regulation (GDPR).

## Acknowledgements

This project was conducted with data and samples from the Berlin Longterm Obsrvation of Vascular Events (BeLOVE) cohort study. The BeLOVE study is substantially funded by the Berlin Institute of Health at Charite (BIH). We thank all participants who took part in the BeLOVE study and the staff in this research program. Study data were collected and managed using REDCap electronic data capture tools hosted at Charité – Universitätsmedizin Berlin^51^. REDCap (Research Electronic Data Capture) is a secure, web-based software platform designed to support data capture for research studies, providing 1) an intuitive interface for validated data capture; 2) audit trails for tracking data manipulation and export procedures; 3) automated export procedures for seamless data downloads to common statistical packages; and 4) procedures for data integration and interoperability with external sources.

## Authors contribution

H.F.A. conceived the project along with K.V., performed data processing, statistical analyses along with M.G.K., interpreted the results, and drafted the first version of the manuscript. S.K.P. contributed to statistical analysis planning and interpretation. MRI preprocessing and technical support were provided by R.M., A.D., S.H., and K.V. Imaging and methodological supervision were performed by T.L., R.M., A.D., C.S., and N.W. Prospective data collection was supported by J.E.W., M.A., K.S., and I.R.N. Project conceptualization and scientific oversight were provided by J.S.M., A.H., M.E., and K.V. All authors discussed the results, contributed to manuscript revisions, and approved the final version of the manuscript.

## BeLOVE Study Group

Ahmadi M, Boldt LH, Buchmann N, Eckardt KU, Edelmann F, Endres M, Gerhardt H, Grittner U, Hübner N, Heege J, Landmesser U, Mai K, Müller DN, Nolte CH, Pinto RM, Piper SK, Pischon T, Rattan S, Rohrpasser-Napierkowski, Schönrath K, Schulz-Menger J, Schweizerhof O, Weber JE.

## Abbreviations

CSVD: Cerebral Small Vessel Disease
WMH: White Matter Hyperintensities
NAWM: Normal-appearing White Matter
pNAWM: perilesional Normal-Appearing White Matter
CVRF: Cerebro-Vascular Risk Factors
DM: Diabetes Mellitus
qMRI: Quantitative Magnetic Resonance Imaging
MPM: Multiparametric Mapping
MTsat: Magnetization Transfer saturation map
R1: Longitudinal Relaxation Rate map
PD: Proton Density map
cWM: contralesional White Matter
BeLOVE: Berlin Longterm Observational of Vascular Events study
ARWMC: Age-Related White Matter Changes scale
FLAIR: FLuid-Attenuated Inversion Recovery sequence

