## Supplemental Materials for "Microstructural Alterations in White Matter Hyperintensities and Perilesional Normal-Appearing White Matter Assessed by Quantitative Multiparametric Mapping - A BeLOVE Study"

Corresponding author: Huma Fatima Ali

**Supplementary Table 1:** Descriptive Statistics of the separate BeLOVE diagnostic arms

|  |  | Acute Kidney Failure | Diabetes | Myocardial Infarction | Acute Heart Failure | Stroke |
| --- | --- | --- | --- | --- | --- | --- |
|  |  | N = 04 | N = 61 | N = 42 | N = 01 | N = 137 |
| Age | Mean (±SD) | 59 (18) | 59 (12) | 64 (11) | 76 (NA) | 63 (11) |
| Sex | Male, n(%) | 2 (50.0) | 38 (62.3) | 31 (73.8) | 1 (100.0) | 100 (73.0) |
|  | Female, n(%) | 2 (50.0) | 23 (37.7) | 11 (26.2) | 0 (0.0) | 37 (27.0) |
| Smoking | Yes, n(%) | 0 (0.0) | 13 (21.3) | 11 (26.2) | 0 (0.0) | 28 (21.7) |
|  | No, n(%) | 0 (0.0) | 37 (60.7) | 17 (40.5) | 0 (0.0) | 66 (51.2) |
|  | Ex-smoker, n(%) | 3 (100.0) | 11 (18.0) | 14 (33.3) | 1 (100.0) | 35 (27.1) |
| Pack years | Median (IQR) | NA | 14  (5-35) | 21.8  (12.3-42.8) | NA | 23.8  (14.1-41.8) |
| Serum Creatinine (mg/dL) | Median (IQR) | 1.9  (1.8-2.6) | 0.88  (0.75-1.05) | 0.9  (0.8-1.1) | 1.13 | 0.9  (0.7-1) |
| Estimated GFR (CKD-EPI, mL/min/1.73m^2^) | Median (IQR) | 27  (23.5-32.5) | 86  (65-90) | 72.5  (67-81.5) | 63 | 83  (74-90) |
| Troponin (ng/L) | Median (IQR) | 43  (30-69) | 8.5  (7-13.8) | 1143  (220-1698.5) | 73 | 9  (7-13) |
| NT-proBNP (pg/mL) | Median (IQR) | 1357  (820.5-3726) | 52  (24.5-112.5) | 474  (232.5-1118.5) | 1770 | 81  (36-249.5) |
| MR-proANP (pmol/L) | Median (IQR) | 511  (504.5-630) | 56  (40.5-92.5) | 94  (77-124) | 425 | 79  (52-111) |
| Interleukin-6 (pg/mL) | Median (IQR) | 29.8  (19.2-30.1) | 2.2  (1.5-3.4) | 5  (3.3-11.6) | 15.1 | 3.7  (2.5-5.6) |
| C-reactive protein (mg/L) | Median (IQR) | 10.7  (8.2-21.4) | 1.5  (0.7-3.1) | 6.8  (2.3-17.4) | 3.3 | 1.6  (0.7-4.6) |
| HbA1c (%) | Median (IQR) | 5.1  (4.8-6.2) | 7.1  (6.6-8.2) | 5.7  (5.3-5.9) | 5.9 | 5.5  (5.3-5.8) |
| Lipoprotein (a) (mg/dL) | Median (IQR) | 3.5  (3.5-8.8) | 7  (7-31.3) | 35.75  (16.7-147.4) | 13.2 | 21.8  (11.9-55.2) |
| Total Cholesterol (mg/dL) | Median (IQR) | 110  (93-127.5) | 162  (137.3-190.3) | 166  (149-194) | 99 | 175  (158-210) |
| HDL Cholesterol (mg/dL) | Median (IQR) | 30  (27-32) | 50  (38.3-61.8) | 50  (39.5-56) | 43 | 50  (42-63) |
| LDL Cholesterol (mg/dL) | Median (IQR) | 67  (54.5-81) | 89  (71-123.3) | 102  (87.5-133.5) | 56 | 111  (87-138.5) |
| Albumin-to-creatinine ratio (urine, mg/g) | Median (IQR) | 19  (13.5-24.5) | 11  (5-32.8) | 6  (5-15.5) | NA | 5.5  (4-14) |
| Fasting Insulin (µU/mL) | Median (IQR) | 13.9  (9.51-33) | 15.5  (8.1-23.5) | 9.4  (7-14.8) | 6.9 | 11.4  (6.9-17.6) |
| R-HOMA Index at day 90 | Median (IQR) | 2.8  (2.05-12.1) | 5.2  (3.5-9) | 2.2  (1.7-3.1) | 1.5 | 2.8  (1.5-4.7) |
| Any cardiovascular medication | Yes, n(%) | 3 (75.0) | 57 (96.6) | 42 (100.0) | 1 (100.0) | 136 (99.3) |
|  | No, n(%) | 0 (0.0) | 1 (1.7) | 0 (0.0) | 0 (0.0) | 1 (0.7) |
|  | Unnamed, n(%) | 1 (25.0) | 1 (1.7) | 0 (0.0) | 0 (0.0) | 0 (0.0) |
| Antihypertensive medication | Yes, n(%) | 3 (100.0) | 43 (75.4) | 42 (100.0) | 1 (100.0) | 86 (63.7) |
|  | No, n(%) | 0 (0.0) | 14 (24.6) | 0 (0.0) | 0 (0.0) | 48 (35.6) |
|  | Unnamed, n(%) | 0 (0.0) | 0 (0.0) | 0 (0.0) | 0 (0.0) | 1 (0.7) |
| Antidiabetic medication | Yes, n(%) | 0 (0.0) | 53 (93.0) | 3 (7.1) | 0 (0.0) | 15 (11.1) |
|  | No, n(%) | 3 (100.0) | 4 (7.0) | 38 (90.5) | 1 (100.0) | 119 (88.1) |
|  | Unnamed, n(%) | 0 (0.0) | 0 (0.0) | 1 (2.4) | 0 (0.0) | 1 (0.7) |
| Lipid lowering medication | Yes, n(%) | 1 (33.3) | 40 (70.2) | 39 (92.9) | 1 (100.0) | 113 (83.1) |
|  | No, n(%) | 2 (66.7) | 17 (29.8) | 3 (7.1) | 0 (0.0) | 23 (16.9) |
| History of Heart Failure* | Yes, n(%) | 0 (0.0) | 5 (8.3) | 1 (2.4) | 1 (100.0) | 3 (2.4) |
|  | No, n(%) | 3 (100.0) | 54 (90.0) | 41 (97.6) | 0 (0.0) | 120 (95.2) |
|  | Unknown, n(%) | 0 (0.0) | 1 (1.7) | 0 (0.0) | 0 (0.0) | 3 (2.4) |
| History of Myocardial infarction* | Yes, n(%) | 0 (0.0) | 4 (6.7) | 7 (16.7) | 0 (0.0) | 4 (3.1) |
|  | No, n(%) | 3 (100.0) | 54 (90.0) | 35 (83.3) | 0 (0.0) | 121 (95.3) |
|  | Unknown, n(%) | 0 (0.0) | 2 (3.3) | 0 (0.0) | 1 (100.0) | 2 (1.6) |
| Hyperlipidemia* | Yes, n(%) | 1 (25.0) | 7 (11.7) | 3 (7.1) | 1 (100.0) | 24 (17.5) |
|  | No, n(%) | 3 (75.0) | 53 (88.3) | 39 (92.9) | 0 (0.0) | 113 (82.5) |
| Atrial fibrillation* | Yes, n(%) | 1 (25.0) | 1 (1.7) | 3 (7.1) | 1 (100.0) | 17 (12.4) |
|  | No, n(%) | 3 (75.0) | 59 (98.3) | 39 (92.9) | 0 (0.0) | 120 (87.6) |
| NIHSS at admission | Mean (±SD) | NA | NA | NA | NA | 2.2 (4.0) |
| mRS at discharge | Mean (±SD) | NA | NA | NA | NA | 0.7 (1.6) |
| TOAST classification | Other etiology | 0 | 0 | 0 | 0 | 4 (3.9) |
|  | Cardioembolic | 0 | 0 | 0 | 0 | 23 (22.3) |
|  | Large vessel angiopathy | 0 | 0 | 0 | 0 | 11 (10.7) |
|  | Small vessel angiopathy | 0 | 0 | 0 | 0 | 11 (10.7) |
|  | Undetermined etiology | 0 | 0 | 0 | 0 | 54 (52.4) |
| MoCA Score at baseline | Median (IQR) | 25 (23-25) | 25 (23-25) | 25 (23-25) | NA | 26 (24-28) |
| MoCA Score at follow-up | Median (IQR) | 28 (26-29) | 25 (23-25) | 25 (23-25) | NA | 26 (24-28) |
| Systolic blood pressure (right arm, mmHg) | Mean (±SD) | 118.9 (7.7) | 131.6 (14.4) | 128.8 (18.1) | 84.3 (NA) | 138.1 (19.3) |
| Diastolic blood pressure (right arm, mmHg) | Mean (±SD) | 71.7 (9.3) | 80.3 (8.5) | 75.2 (10.1) | 51.0 (NA) | 81.3 (10.2) |
| Systolic blood pressure (left arm, mmHg) | Mean (±SD) | 120.9 (12.5) | 130.3 (13.8) | 127.0 (18.2) | 83.7 (NA) | 136.6 (17.7) |
| Diastolic blood pressure (left arm, mmHg) | Mean (±SD) | 71.4 (9.1) | 81.2 (9.4) | 75.1 (10.7) | 49.0 (NA) | 80.7 (9.5) |
| Body Mass Index (BMI, kg/m^2^) | Mean (±SD) | 26 (2.4) | 31 (5.8) | 27 (4.2) | 33 (NA) | 26 (4.2) |
| Waist-to-hip ratio | Mean (±SD) | 0.9 (0.1) | 1.0 (0.1) | 1.0 (0.1) | 1.0 (NA) | 1.0 (0.1) |
| Diabetes Mellitus* | No | 2 (50.0) | 0 (0.0) | 31 (75.6) | 1 (100.0) | 110 (81.5) |
|  | Yes | 2 (50.0) | 61 (100.0) | 10 (24.4) | 0 (0.0) | 25 (18.5) |
| Arterial Hypertension* | No | 1 (25.0) | 11 (18.0) | 2 (4.9) | 0 (0.0) | 30 (22.2) |
|  | Yes | 3 (75.0) | 50 (82.0) | 39 (95.1) | 1 (100.0) | 105 (77.8) |
| Intima-media thickness right (mm) | Mean (±SD) | 0.6 (0.2) | 0.7 (0.2) | 0.7 (0.2) | 0.7 (NA) | 0.7 (0.2) |
| Intima-media thickness left (mm) | Mean (±SD) | 0.7 (0.3) | 0.7 (0.2) | 0.7 (0.2) | 0.7 (NA) | 0.7 (0.2) |
| ARWMC Score | Median (IQR) | 3 (1-2) | 3 (1-2) | 3 (1-2) | 2 (NA) | 4 (2-3) |
| ARWMC Group | Mild lesion load | 3 (75.0) | 44 (72.1) | 27 (65.9) | 1 (100.0) | 76 (54.7) |
|  | Moderate lesion load | 1 (25.0) | 17 (27.9) | 14 (34.1) | 0 (0.0) | 63 (45.3) |
| MTsat in cWM | Mean (±SD) | 3.5 (0.9) | 3.7 (0.4) | 3.5 (0.4) | 3.0 (NA) | 3.6 (0.5) |
| MTsat in WMH | Mean (±SD) | 3.3 (0.5) | 3.2 (0.4) | 3.0 (0.4) | 2.7 (NA) | 3.1 (0.4) |
| MTsat in pNAWM at 1mm | Mean (±SD) | 3.6 (0.4) | 3.5 (0.4) | 3.4 (0.3) | 2.9 (NA) | 3.4 (0.4) |
| MTsat in pNAWM at 2mm | Mean (±SD) | 3.7 (0.4) | 3.5 (0.4) | 3.4 (0.3) | 3.2 (NA) | 3.5 (0.4) |
| MTsat in pNAWM at 3mm | Mean (±SD) | 3.6 (0.5) | 3.3 (0.4) | 3.3 (0.3) | 3.2 (NA) | 3.5 (0.4) |
| R1 in cWM | Mean (±SD) | 1.0 (0.2) | 0.9 (0.1) | 0.9 (0.1) | 0.9 (NA) | 1.0 (0.4) |
| R1 in WMH | Mean (±SD) | 1.0 (0.1) | 0.8 (0.1) | 0.9 (0.1) | 0.8 (NA) | 0.9 (0.3) |
| R1 in pNAWM at 1mm | Mean (±SD) | 1.0 (0.1) | 0.9 (0.1) | 0.9 (0.1) | 0.9 (NA) | 1.0 (0.3) |
| R1 in pNAWM at 2mm |  | 1.0 (0.1) | 0.9 (0.1) | 0.9 (0.1) | 0.9 (NA) | 1.0 (0.3) |
| R1 in pNAWM at 3mm | Mean (±SD) | 1.0 (0.1) | 0.9 (0.1) | 0.9 (0.1) | 0.9 (NA) | 1.0 (0.3) |
| PD in cWM | Mean (±SD) | 70.6 (4.6) | 69.3 (1.9) | 70.0 (2.5) | 70.2 (NA) | 69.3 (3.0) |
| PD in WMH | Mean (±SD) | 71.2 (2.6) | 72.1 (2.2) | 72.5 (2.4) | 73.9 (NA) | 71.4 (2.7) |
| PD in pNAWM at 1mm | Mean (±SD) | 70.6 (1.9) | 70.6 (1.7) | 70.6 (1.7) | 87.4 (NA) | 69.8 (2.2) |
| PD in pNAWM at 2mm | Mean (±SD) | 70.2 (1.6) | 70.6 (1.6) | 70.2 (1.8) | 69.5 (NA) | 69.5 (2.1) |
| PD in pNAWM at 3mm | Mean (±SD) | 70.5 (1.7) | 71.1 (1.6) | 70.6 (2.0) | 69.3 (NA) | 70.0 (2.3) |

GFR = Glomerular Filtration Rate; HDL = High-Density Lipoprotein; LDL = Low-Density Lipoprotein; R-HOMA = Revised-Homeostatic Model Assessement ; NIHSS = National Institute of Health Stroke Scale; TOAST = Trial of ORG 10172 in Acute Stroke Treatment ; MoCA =Montreal Cognitive Assessment; ARWMC = Age-Related White Matter Changes score; WMH = White Matter Hyperintensities; cWM =contralesional White Matter; pNAWM = perilesional Normal-Appearing White Matter; IQR =Inter-Quartile Range; SD = Standard Deviation.

*The presence of diabetes and arterial hypertension was defined by patient´s self-reported history, reports in medical records, current medication or current measures of glucose metabolism or blood pressure. Smoking status were evaluated by self-report and medical records, while information on the presence of hyperlipidemia, atrial fibrillation and coronary artery disease was based on medical records only and historical Information on prior heart failure and myocardial infarction was taken from self-report.

**Supplementary Table 2a**: Linear mixed effects models demonstrating association of CVRF with MPM metrics and their corresponding perilesional NAWM of 1, 2, and 3 mm (MPM ~ CVRF + region+CVRF*region +(1|ID).

| CVRF | MPM Map | Region | ß (95% CI) | p-value |
| --- | --- | --- | --- | --- |
| Age (n = 245) | MTsat | Main effect | -0.015 (-0.019 – -0.011) | <0.001 |
|  |  | pNAWM at 1mm | 0.001 (-0.001 – 0.004) | 0.421 |
|  |  | pNAWM at 2mm | 0.002 (-0.000 - 0.005) | 0.062 |
|  |  | pNAWM at 3mm | 0.003 (0.000 - 0.006) | 0.008 |
|  | R1 | Main effect | -0.002 (-0.005 – 0.000) | 0.064 |
|  |  | pNAWM at 1mm | 0.000 (-0.001 -0.000) | 0.300 |
|  |  | pNAWM at 2mm | 0.000 (-0.000 – 0.000) | 0.155 |
|  |  | pNAWM at 3mm | 0.000 (-0.000 – 0.000) | 0.159 |
|  | PD | Main effect | -0.002 (-0.031 – 0.025) | 0.839 |
|  |  | pNAWM at 1mm | 0.005 (-0.014 – 0.025) | 0.582 |
|  |  | pNAWM at 2mm | -0.007 (-0.027 – 0.012) | 0.443 |
|  |  | pNAWM at 3mm | -0.013 (-0.034 – 0.006) | 0.174 |
| Sex (female)  (n=73) | MTsat | Main effect | -0.021 (-0.131 – 0.087) | 0.697 |
|  |  | pNAWM at 1mm | -0.018 (-0.085 – 0.047) | 0.579 |
|  |  | pNAWM at 2mm | -0.033 (-0.099 - -0.033) | 0.334 |
|  |  | pNAWM at 3mm | -0.035 (-0.103 – -0.031) | 0.296 |
|  | R1 | Main effect | 0.051 (-0.015 – 0.117) | 0.132 |
|  |  | pNAWM at 1mm | 0.008 (-0.005 – 0.022) | 0.234 |
|  |  | pNAWM at 2mm | 0.010 (-0.003 – 0.023) | 0.156 |
|  |  | pNAWM at 3mm | 0.009 (-0.004 – 0.024) | 0.162 |
|  | PD | Main effect | -0.055 (-0.632 – 0.742) | 0.876 |
|  |  | pNAWM at 1mm | -0.017 (-0.500 – 0.465) | 0.943 |
|  |  | pNAWM at 2mm | 0.156 (-0.327 – 0.639) | 0.525 |
|  |  | pNAWM at 3mm | 0.222 (-0.263 – 0.708) | 0.369 |
| Diabetes Mellitus  (n=98) | MTsat | Main effect | 0.021 (-0.081 – 0.123) | 0.685 |
|  |  | pNAWM at 1mm | -0.019 (-0.081 – 0.042) | 0.533 |
|  |  | pNAWM at 2mm | -0.067 (-0.128 – -0.005) | 0.034 |
|  |  | pNAWM at 3mm | -0.123 (-0.185 – -0.061) | 1.138 |
|  | R1 | Main effect | -0.059 (-0.122 – 0.002) | 0.057 |
|  |  | pNAWM at 1mm | -0.007 (-0.020 – 0.005) | 0.285 |
|  |  | pNAWM at 2mm | -0.011 (-0.024 – 0.001) | 0.088 |
|  |  | pNAWM at 3mm | -0.017 (-0.031 – -0.004) | 0.009 |
|  | PD | Main effect | 0.573 (-0.088 – 1.233) | 0.089 |
|  |  | pNAWM at 1mm | -0.197 (-0.664 – 0.269) | 0.406 |
|  |  | pNAWM at 2mm | 0.037 (-0.430 – 0.504) | 0.876 |
|  |  | pNAWM at 3mm | 0.077 (-0.391 – 0.546) | 0.745 |
| Arterial Hypertension  (n=198) | MTsat | Main effect | -0.018 (-0.151 – 0.115) | 0.791 |
|  |  | pNAWM at 1mm | -0.044 (-0.126 – 0.037) | 0.280 |
|  |  | pNAWM at 2mm | 0.033 (-0.114 – 0.049) | 0.433 |
|  |  | pNAWM at 3mm | 0.039 (-0.121 – 0.043) | 0.353 |
|  | R1 | Main effect | -0.059 (-0.139 – 0.021) | 0.145 |
|  |  | pNAWM at 1mm | -0.001 (-0.018 – 0.015) | 0.861 |
|  |  | pNAWM at 2mm | 0.008 (-0.008 – 0.025) | 0.337 |
|  |  | pNAWM at 3mm | 0.011 (-0.005 – 0.028) | 0.180 |
|  | PD | Main effect | 0.489 (.0.347 – 1.327) | 0.251 |
|  |  | pNAWM at 1mm | 0.026 (-0.563 – 0.616) | 0.929 |
|  |  | pNAWM at 2mm | -0.148 (-0.738 – 0.441) | 0.623 |
|  |  | pNAWM at 3mm | -0.108 (-0.703 – 0.486) | 0.720 |
| Hyperlipiproteinemia  (n=215) | MTsat | Main effect | -0.055 (-0.196 – 0.085) | 0.441 |
|  |  | pNAWM at 1mm | 0.023 (-0.062 – 0.109) | 0.588 |
|  |  | pNAWM at 2mm | 0.069 (-0.017 – 0.155) | 0.114 |
|  |  | pNAWM at 3mm | 0.083 (-0.003 – 0.169) | 0.058 |
|  | R1 | Main effect | 0.032 (-0.054 – 0.119) | 0.461 |
|  |  | pNAWM at 1mm | 0.012 (-0.005 – 0.030) | 0.175 |
|  |  | pNAWM at 2mm | 0.024 (0.005 – 0.042) | 0.010 |
|  |  | pNAWM at 3mm | 0.023 (0.004 – 0.041) | 0.013 |
|  | PD | Main effect | -1.217 (-2.098 – -0.337) | 0.007 |
|  |  | pNAWM at 1mm | 0.741 (0.118 – 1.364) | 0.019 |
|  |  | pNAWM at 2mm | 0.297 (-0.325 – 0.920) | 0.348 |
|  |  | pNAWM at 3mm | 0.229 (-0.394 – 0.852) | 0.471 |
| Coronary Artery Disease  (n=60) | MTsat | Main effect | -0.095 (-0.210 - 0.019) | 0.102 |
|  |  | pNAWM at 1mm | -0.007 (-0.077 – 0.063) | 0.836 |
|  |  | pNAWM at 2mm | -0.000 (-0.071 – 0.069) | 0.992 |
|  |  | pNAWM at 3mm | -0.019 (-0.090 - 0.052) | 0.595 |
|  | R1 | Main effect | 0.011 (-0.059 – 0.082) | 0.750 |
|  |  | pNAWM at 1mm | 0.007 (-0.008 – 0.023) | 0.355 |
|  |  | pNAWM at 2mm | 0.011 (-0.003 – 0.026) | 0.136 |
|  |  | pNAWM at 3mm | 0.010 (-0.004 – 0.025) | 0.171 |
|  | PD | Main effect | 0.806 (0.102 – 1.509) | 0.025 |
|  |  | pNAWM at 1mm | 0.160 (-0.335 – 0.656) | 0.526 |
|  |  | pNAWM at 2mm | -0.369 (-0.864 – 0.126) | 0.144 |
|  |  | pNAWM at 3mm | -0.467 (-0.966 – 0.031) | 0.066 |
| Atrial Fibrillation  (n=25) | MTsat | Main effect | -0.154 (-0.327 – 0.018) | 0.081 |
|  |  | pNAWM at 1mm | 0.033 (-0.073 – 0.139) | 0.539 |
|  |  | pNAWM at 2mm | 0.042 (-0.064 – 0.138) | 0.539 |
|  |  | pNAWM at 3mm | 0.046 (-0.059 – 0.152) | 0.391 |
|  | R1 | Main effect | -0.050 (-0.153 – 0.053) | 0.339 |
|  |  | pNAWM at 1mm | 0.007 (-0.015 – 0.028) | 0.540 |
|  |  | pNAWM at 2mm | 0.007 (-0.014 – 0.029) | 0.491 |
|  |  | pNAWM at 3mm | 0.008 (-0.013 – 0.030) | 0.437 |
|  | PD | Main effect | -0.193 (-1.229 – 0.843) | 0.714 |
|  |  | pNAWM at 1mm | 0.964 (0.239 – 1.688) | 0.009 |
|  |  | pNAWM at 2mm | 0.122 (-0.602 -0.847) | 0.739 |
|  |  | pNAWM at 3mm | 0.161 (-0.564 – 0.885) | 0.663 |
| Smoking (Current smoker)  (n=52) | MTsat | Main effect | 0.139 (0.001 – 0.276) | 0.048 |
|  |  | pNAWM at 1mm | 0.008 (-0.075 – 0.093) | 0.839 |
|  |  | pNAMM at 2mm | 0.003 (-0.081 – 0.087) | 0.937 |
|  |  | pNAWM at 3mm | -0.012 (-0.097 – 0.072) | 0.766 |
|  | R1 | Main effect | 0.076 (-0.007 – 0.160) | 0.072 |
|  |  | pNAWM at 1mm | 0.008 (-0.009 – 0.025) | 0.362 |
|  |  | pNAWM at 2mm | 0.009 (-0.008 – 0.026) | 0.311 |
|  |  | pNAWM at 3mm | 0.009 (-0.008 – 0.026) | 0.305 |
|  | PD | Main effect | -0.009 (-0.881 – 0.861) | 0.983 |
|  |  | pNAWM at 1mm | 0.161 (-0.445 – 0.768) | 0.602 |
|  |  | pNAWM at 2mm | 0.258 (-0.349 – 0.865) | 0.404 |
|  |  | pNAWM at 3mm | 0.166 (-0.443 – 0.775) | 0.593 |
| Waist-hip-ratio  (n=243) | MTsat | Main effect | 0.096 (-0.433 – 0.628) | 0.719 |
|  |  | pNAWM at 1mm | -0.121 (-0.443 – 0.201) | 0.462 |
|  |  | pNAWM at 2mm | -0.196 (-0.517 – 0.126) | 0.233 |
|  |  | pNAWM at 3mm | 0.357 (-0.685 – -0.031) | 0.032 |
|  | R1 | Main effect | -0.404 (-0.727 – -0.082) | 0.014 |
|  |  | pNAWM at 1mm | -0.076 (-0.144 – -0.008) | 0.027 |
|  |  | pNAWM at 2mm | -0.100 (-0.168 – -0.032) | 0.004 |
|  |  | pNAWM at 3mm | -0.118 (-0.187 – -0.049) | <0.001 |
|  | PD | Main effect | -1.047 (-4.485 – 2.390) | 0.549 |
|  |  | pNAWM at 1mm | -0.080 (-2.496 – 2.336) | 0.947 |
|  |  | pNAWM at 2mm | 0.487 (-1.929 – 2.903) | 0.692 |
|  |  | pNAWM at 3mm | 0.461 (0.002 – 0.022) | 0.709 |
| Body Mass Index  (N=243) | MTsat | Main effect | 0.012 (0.002 – 0.022) | 0.016 |
|  |  | pNAWM at 1mm | -0.003 (-0.009 – 0.002) | 0.233 |
|  |  | pNAWM at 2mm | -0.007 (-0.013 – -0.000) | 0.023 |
|  |  | pNAWM at 3mm | -0.009 (-0.015 - - 0.003) | 0.002 |
|  | R1 | Main effect | -0.005 (-0.012 - -0.000) | 0.068 |
|  |  | pNAWM at 1mm | -0.002 (-0.003 – -0.000) | 0.004 |
|  |  | pNAWM at 2mm | -0.002 (-0.003 – -0.000) | 0.001 |
|  |  | pNAWM at 3mm | -0.002 (-0.003 – -0.000) | 0.005 |
|  | PD | Main effect | 0.007 (-0.057 – 0.071) | 0.829 |
|  |  | pNAWM at 1mm | 0.023 (-0.022 – 0.068) | 0.324 |
|  |  | pNAWM at 2mm | 0.018 (-0.027 – 0.064) | 0.427 |
|  |  | pNAWM at 3mm | 0.018 (-0.026 – 0.064) | 0.418 |
| C-Reactive Protein (CRP)  (n=224) | MTsat | Main effect | 0.012 (0.000 – 0.024) | 0.038 |
|  |  | pNAWM at 1mm | -0.004 (-0.011 – 0.002) | 0.213 |
|  |  | pNAWM at 2mm | -0.005 (-0.012 – 0.001) | 0.121 |
|  |  | pNAWM at 3mm | -0.006 (-0.013 – 0.000) | 0.087 |
|  | R1 | Main effect | -0.000 (-0.004 – 0.002) | 0.524 |
|  |  | pNAWM at 1mm | 0.000 (-0.000 – 0.000) | 0.792 |
|  |  | pNAWM at 2mm | 0.000 (-0.000 - 0.000) | 0.699 |
|  |  | pNAWM at 3mm | 0.000 (-0.000 – 0.001) | 0.495 |
|  | PD | Main effect | -0.002 (-0.039 – 0.035) | 0.911 |
|  |  | pNAWM at 1mm | -0.005 (-0.031 – 0.021) | 0.698 |
|  |  | pNAWM at 2mm | -0.006 (-0.033 – 0.019) | 0.625 |
|  |  | pNAWM at 3mm | -0.009 (-0.035 – 0.017) | 0.495 |
| Alpha Lipoprotein  (n=215) | MTsat | Main effect | -0.000 (-0.000 – 0.000) | 0.256 |
|  |  | pNAWM at 1mm | -0.000 (-0.000 – 0.000) | 0.948 |
|  |  | pNAWM at 2mm | 0.000 (-0.000 – 0.000) | 0.663 |
|  |  | pNAWM at 3mm | 0.000 (-0.000 – 0.000) | 0.593 |
|  | R1 | Main effect | -0.000 (-0.000 – 0.000) | 0.507 |
|  |  | pNAWM at 1mm | -0.000 (-0.000 – 0.000) | 0.229 |
|  |  | pNAWM at 2mm | -0.000 (-0.000 – 0.000) | 0.304 |
|  |  | pNAWM at 3mm | -0.000 (-0.000 – 0.000) | 0.720 |
|  | PD | Main effect | -0.000 (-0.003 – 0.004) | 0.120 |
|  |  | pNAWM at 1mm | -0.002 (-0.004 – 0.000) | 0.215 |
|  |  | pNAWM at 2mm | -0.002 (-0.004 – 0.000) | 0.169 |
|  |  | pNAWM at 3mm | -0.002 (-0.005 – 0.000) | 0.169 |
| Interleukin  (n=170) | MTsat | Main effect | -0.002 (-0.019 – 0.014) | 0.774 |
|  |  | pNAWM at 1mm | -0.002 (-0.011 – 0.007) | 0.736 |
|  |  | pNAWM at 2mm | -0.001 (-0.011 – 0.008) | 0.782 |
|  |  | pNAWM at 3mm | -0.002 (-0.012 – 0.007) | 0.638 |
|  | R1 | Maine effect | -0.001 (-0.005 – 0.002) | 0.353 |
|  |  | pNAWM at 1mm | 0.000 (-0.000 – 0.002) | 0.458 |
|  |  | pNAWM at 2mm | 0.000 (-0.001 – 0.002) | 0.535 |
|  |  | pNAWM at 3mm | 0.000 (-0.001 – 0.001) | 0.858 |
|  | PD | Main effect | -0.019 (-0.113 – 0.073) | 0.675 |
|  |  | pNAWM at 1mm | 0.021 (-0.044 – 0.086) | 0.534 |
|  |  | pNAWM at 2mm | -0.021 (-0.086 – 0.044) | 0.528 |
|  |  | pNAWM at 3mm | -0.016 (-0.081 - 0.049) | 0.627 |
| Albumin-Creatinine ratio in urine  (n=170) | MTsat | Main effect | -0.000 (-0.00 – 0.000) | 0.796 |
|  |  | pNAWM at 1mm | -0.000 (-0.00 – 0.000) | 0.824 |
|  |  | pNAWM at 2mm | 0.000 (-0.00 – 0.000) | 0.930 |
|  |  | pNAWM at 3mm | 0.000 (-0.00 – 0.000) | 0.845 |
|  | R1 | Main effect | -0.000 (-0.000 - 0.000) | 0.798 |
|  |  | pNAWM at 1mm | 0.000 (-0.000 - 0.000) | 0.838 |
|  |  | pNAWM at 2mm | 0.000 (-0.000 - 0.000) | 0.607 |
|  |  | pNAWM at 3mm | 0.000 (-0.000 - 0.000) | 0.381 |
|  | PD | Main effect | -0.000 (-0.000 – 0.000) | 0.987 |
|  |  | pNAWM at 1mm | 0.000 (-0.000 – 0.000) | 0.970 |
|  |  | pNAWM at 2mm | -0.000 (-0.000 – 0.000) | 0.738 |
|  |  | pNAWM at 3mm | -0.000 (-0.000 – 0.000) | 0.815 |
| R-HOMA Index at day 90  (n=195) | MTsat | Main effect | 0.011 (-0.002 – 0.024) | 0.099 |
|  |  | pNAWM at 1mm | -0.004 (-0.013 – 0.003) | 0.273 |
|  |  | pNAWM at 2mm | -0.010 (-0.018 – -0.002) | 0.015 |
|  |  | pNAWM at 3mm | -0.017 (-0.026 - -0.009) | <0.001 |
|  | R1 | Main effect | -0.002 (-0.009 – 0.004) | 0.459 |
|  |  | pNAWM at 1mm | -0.001 (-0.003 – 0.000) | 0.079 |
|  |  | pNAWM at 2mm | -0.002 (-0.004 – -0.000) | 0.013 |
|  |  | pNAWM at 3mm | -0.003 (-0.004 – -0.000) | 0.003 |
|  | PD | Main effect | 0.002 (-0.088 – 0.093) | 0.962 |
|  |  | pNAWM at 1mm | -0.002 (-0.066 – 0.061) | 0.948 |
|  |  | pNAWM at 2mm | 0.015 (-0.048 – 0.078) | 0.650 |
|  |  | pNAWM at 3mm | 0.022 (-0.041 – 0.085) | 0.493 |
| Carotid Intima-media thickness  (n=222) | MTsat | Main effect | -0.334 (-0.693 - 0.025) | 0.068 |
|  |  | pNAWM at 1mm | 0.021 (-0.202 – 0.245) | 0.852 |
|  |  | pNAWM at 2mm | 0.085 (-0.138 – 0.309) | 0.454 |
|  |  | pNAWM at 3mm | 0.116 (-0.111 – 0.342) | 0.316 |
|  | R1 | Main effect | -0.115 (-0.281 – 0.051) | 0.173 |
|  |  | pNAWM at 1mm | -0.014 (-0.054 – 0.026) | 0.506 |
|  |  | pNAWM at 2mm | -0.008 (-0.048 – 0.032) | 0.694 |
|  |  | pNAWM at 3mm | 0.004 (-1.591 – 3.203) | 0.858 |
|  | PD | Main effect | 0.806 (-1.591 – 3.203) | 0.508 |
|  |  | pNAWM at 1mm | -0.347 (-2.051 – 1.355) | 0.689 |
|  |  | pNAWM at 2mm | -0.822 (-2.525 – 0.881) | 0.343 |
|  |  | pNAWM at 3mm | -1.376 (-3.089 – 0.336) | 0.115 |
| Pulse Wave velocity  (n=202) | MTsat | Main effect | -0.015 (-0.039 – 0.010) | 0.249 |
|  |  | pNAWM at 1mm | -0.002 (-0.017 – 0.013) | 0.765 |
|  |  | pNAWM at 2mm | -0.002 (-0.017 – 0.012) | 0.751 |
|  |  | pNAWM at 3mm | -0.006 (-0.022 – 0.008) | 0.413 |
|  | R1 | Main effect | -0.008 (-0.021 – 0.003) | 0.151 |
|  |  | pNAWM at 1mm | -0.002 (-0.005 – 0.001) | 0.279 |
|  |  | pNAWM at 2mm | -0.002 (-0.005 – 0.001) | 0.207 |
|  |  | pNAWM at 3mm | -0.003 (0.005 – 0.000) | 0.087 |
|  | PD | Main effect | -0.146 (-0.301 – 0.008) | 0.064 |
|  |  | pNAWM at 1mm | -0.027 (-0.138 – 0.084) | 0.628 |
|  |  | pNAWM at 2mm | 0.019 (-0.092 – 0.130) | 0.735 |
|  |  | pNAWM at 3mm | 0.013 (-0.098 – 0.125) | 0.815 |

pNAWM = perilesional Normal-Appearing White Matter; R-HOMA = Revised-Homeostatic Model Assessment; CVRF = Cerebrovascular risk factors; MPM =multi-parametric mapping; 95% CI = 95% confidence intervals.

**Supplementary Table 2b**: Sensitivity Analysis excluding patients taking medication: antihypertensive, antidiabetic, and lipid-lowering drugs.

| CVRF | MPM Map | Region | ß (95% CI) | p-value |
| --- | --- | --- | --- | --- |
| Arterial Hypertension | MTsat | Main effect | 0.008 (-1.85 – 1.87) | 0.936 |
|  |  | pNAWM at 1mm | -0.023 (-0.29 – 0.24) | 0.861 |
|  |  | pNAWM at 2mm | 0.001 (-0.26 – 0.26) | 0.991 |
|  |  | pNAWM at 3mm | 0.034 (-0.23 – 0.29) | 0.799 |
|  | R1 | Main effect | 0.051 (-0.20 – 0.30) | 0.693 |
|  |  | pNAWM at 1mm | -0.018 (-0.38 – 0.34) | 0.920 |
|  |  | pNAWM at 2mm | 0.003 (-0.35 – 0.36) | 0.989 |
|  |  | pNAWM at 3mm | 0.009 (-0.35 – 0.37) | 0.960 |
|  | PD | Main effect | 0.637 (-1.25 – 2.53) | 0.509 |
|  |  | pNAWM at 1mm | -0.251 (-2.92 – 0.42) | 0.854 |
|  |  | pNAWM at 2mm | -0.194 (-2.86 – 2.47) | 0.887 |
|  |  | pNAWM at 3mm | -0.206 (-2.91 – 2.49) | 0.882 |
| Diabetes Mellitus | MTsat | Main effect | 0.106 (-0.07 – 0.28) | 0.175 |
|  |  | pNAWM at 1mm | -0.077 (-0.32 – 0.179) | 0.489 |
|  |  | pNAWM at 2mm | -0.077 (-0.32 – 0.17) | 0.501 |
|  |  | pNAWM at 3mm | -0.085 (-0.33 – 0.16) | 0.397 |
|  | R1 | Main effect | -0.051 (-0.18 – 0.08) | 0.491 |
|  |  | pNAWM at 1mm | 0.003 (-0.18 – 0.19) | 0.960 |
|  |  | pNAWM at 2mm | 0.006 (-0.18 – 0.19) | 0.993 |
|  |  | pNAWM at 3mm | 0.009 (-0.18 – 0.20) | 0.958 |
|  | PD | Main effect | 0.361 (0.83 – 1.55) | 0.518 |
|  |  | pNAWM at 1mm | 0.086 (-1.59 – 1.77) | 0.925 |
|  |  | pNAWM at 2mm | 0.130 (-1.55 – 1.81) | 0.891 |
|  |  | pNAWM at 3mm | 0.152 (-1.53 – 1.84) | 0.874 |
| Hyperlipoproteinemia | MTsat | Main effect | 0.467 (-0.21 – 1.41) | 0.176 |
|  |  | pNAWM at 1mm | -0.033 (-0.98 – 0.92) | 0.946 |
|  |  | pNAWM at 2mm | -0.013 (-0.97 – 0.94) | 0.979 |
|  |  | pNAWM at 3mm | 0.011 (-0.94 – 0.96) | 0.982 |
|  | R1 | Main effect | -0.061 (-0.88 – 0.76) | 0.884 |
|  |  | pNAWM at 1mm | -0.073 (-1.24 – 1.09) | 0.902 |
|  |  | pNAWM at 2mm | -0.049 (-1.21 – 1.11) | 0.934 |
|  |  | pNAWM at 3mm | -0.023 (-1.19 – 1-14) | 0.969 |
|  | PD | Main effect | NA | NA |
|  |  | pNAWM at 1mm | NA | NA |
|  |  | pNAWM at 2mm | NA | NA |
|  |  | pNAWM at 3mm | NA | NA |

pNAWM = perilesional Normal-Appearing White Matter; CVRF = Cerebrovascular risk factors; MPM =multi-paraametric mapping; 95% CI = 95% confidence intervals.

**Supplementary Figure**

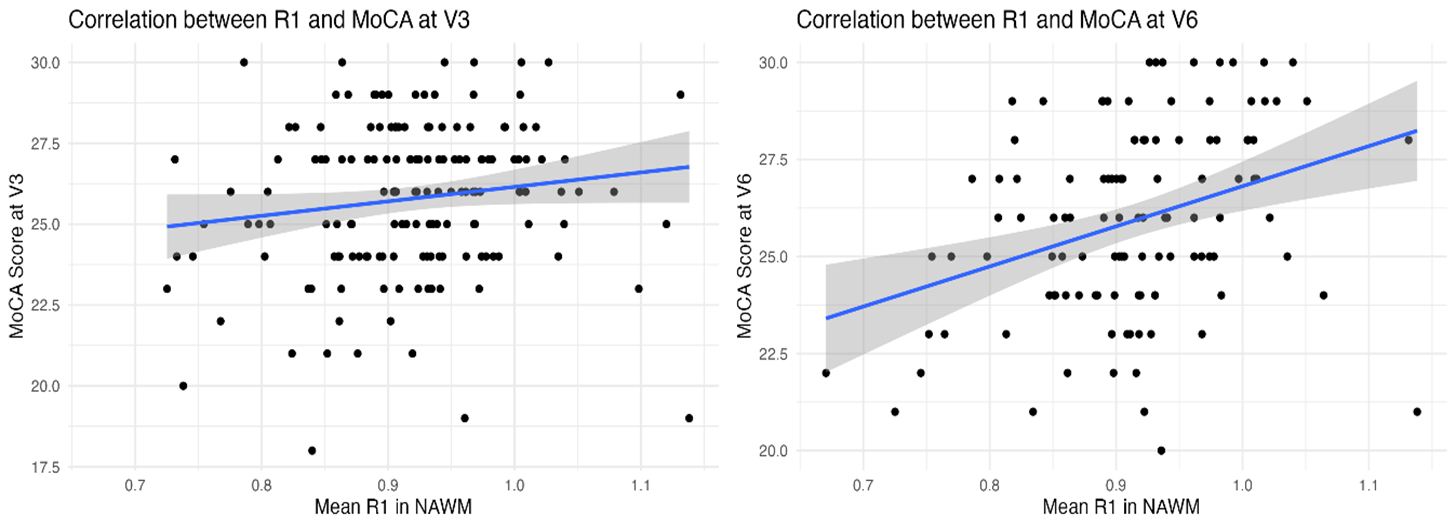

**Supplementary Figure 1:** Correlation plots showing association between mean R1 values in normal-appearing white matter and cognitive performance at baseline (V3) and follow-up (V6).
